# SARS-CoV-2 B.1.1.7 sensitivity to mRNA vaccine-elicited, convalescent and monoclonal antibodies

**DOI:** 10.1101/2021.01.19.21249840

**Authors:** Dami A. Collier, Anna De Marco, Isabella A.T.M. Ferreira, Bo Meng, Rawlings Datir, Alexandra C. Walls, Steven A. Kemp S, Jessica Bassi, Dora Pinto, Chiara Silacci Fregni, Siro Bianchi, M. Alejandra Tortorici, John Bowen, Katja Culap, Stefano Jaconi, Elisabetta Cameroni, Gyorgy Snell, Matteo S. Pizzuto, Alessandra Franzetti Pellanda, Christian Garzoni, Agostino Riva, The CITIID-NIHR BioResource COVID-19 Collaboration, Anne Elmer, Nathalie Kingston, Barbara Graves, Laura E McCoy, Kenneth GC Smith, John R. Bradley, Nigel Temperton, Lourdes Ceron-Gutierrez L, Gabriela Barcenas-Morales, The COVID-19 Genomics UK (COG-UK) consortium, William Harvey, Herbert W. Virgin, Antonio Lanzavecchia, Luca Piccoli, Rainer Doffinger, Mark Wills, David Veesler, Davide Corti, Ravindra K. Gupta

**Affiliations:** Cambridge Institute of Therapeutic Immunology & Infectious Disease (CITIID), Cambridge, UK; Department of Medicine, University of Cambridge, Cambridge, UK; Division of Infection and Immunity, University College London, London, UK; Humabs Biomed SA, a subsidiary of Vir Biotechnology, 6500 Bellinzona, Switzerland; Department of Biochemistry, University of Washington, Seattle, WA 98195, USA; Vir Biotechnology, San Francisco, CA 94158, USA; Clinic of Internal Medicine and Infectious Diseases, Clinica Luganese Moncucco, 6900 Lugano, Switzerland; Division of Infectious Diseases, Luigi Sacco Hospital, University of Milan, Milan, Italy; NIHR Cambridge Clinical Research Facility, Cambridge, UK; NIHR Bioresource, Cambridge, UK; University of Kent, Canturbury, UK; Department of Clinical Biochemistry and Immunology, Addenbrookes Hospital, UK; Laboratorio de Inmunologia, S-Cuautitlán, UNAM, Mexico; Institute of Biodiversity, University of Glasgow, Glasgow, UK; University of KwaZulu Natal, Durban, South Africa; Africa Health Research Institute, Durban, South Africa; Department of Infectious Diseases, Cambridge University Hospitals NHS Trust, Cambridge UK

**Keywords:** SARS-CoV-2, COVID-19, antibody, vaccine, neutralising antibodies, mutation, variant

## Abstract

Severe Acute Respiratory Syndrome Coronavirus-2 (SARS-CoV-2) transmission is uncontrolled in many parts of the world, compounded in some areas by higher transmission potential of the B1.1.7 variant now seen in 50 countries. It is unclear whether responses to SARS-CoV-2 vaccines based on the prototypic strain will be impacted by mutations found in B.1.1.7. Here we assessed immune responses following vaccination with mRNA-based vaccine BNT162b2. We measured neutralising antibody responses following a single immunization using pseudoviruses expressing the wild-type Spike protein or the 8 amino acid mutations found in the B.1.1.7 spike protein. The vaccine sera exhibited a broad range of neutralising titres against the wild-type pseudoviruses that were modestly reduced against B.1.1.7 variant. This reduction was also evident in sera from some convalescent patients. Decreased B.1.1.7 neutralisation was also observed with monoclonal antibodies targeting the N-terminal domain (9 out of 10), the Receptor Binding Motif (RBM) (5 out of 31), but not in neutralising mAbs binding outside the RBM. Introduction of the E484K mutation in a B.1.1.7 background to reflect newly emerging viruses in the UK led to a more substantial loss of neutralising activity by vaccine-elicited antibodies and mAbs (19 out of 31) over that conferred by the B.1.1.7 mutations alone. E484K emergence on a B.1.1.7 background represents a threat to the vaccine BNT162b.

## Introduction

The outbreak of a pneumonia of unknown cause in Wuhan, China in December 2019, culminated in a global pandemic due to a novel viral pathogen, now known to be SARS-CoV-2^1^. The unprecedented scientific response to this global challenge has led to the rapid development of vaccines aimed at preventing SARS-COV-2 infection and transmission. Continued viral evolution led to the emergence and selection of SARS-CoV-2 variants with enhanced infectivity/transmissibility^2,3^ ^4,5^ and ability to circumvent drug^6^ and immune control^7,8^.

SARS-CoV-2 vaccines have recently been licensed that target the spike (S) protein, either using mRNA or adenovirus vector technology with protection rates ranging from 62 to 95%^9-11^. The BNT162b2 vaccine encodes the full-length trimerised S protein of SARS CoV-2 and is formulated in lipid nanoparticles for delivery to cells^12^. Other vaccines include the Moderna mRNA-1273 vaccine, which is also a lipid nanoparticle formulated S glycoprotein^13^ and the Oxford-AstraZeneca ChAdOx1 nCoV-19 vaccine (AZD1222) which is a replication-deficient chimpanzee adenoviral vector ChAdOx1, containing the S glycoprotein^14^. The duration of immunity conferred by these vaccines is as yet unknown. These vaccines were designed against the Wuhan-1 isolate discovered in 2019. Concerns have been raised as to whether these vaccines will be effective against newly emergent SARS-CoV-2 variants, such as B.1.1.7 (N501Y.V1), B.1.351 (N501Y.V2) and P1 (N501Y.V3) that originated in the UK, South Africa, and Brazil and are now being detected all over the world^15-17^.

In clinical studies of the Pfizer-BioNTech BNT162b2 vaccine, high levels of protection against infection and severe disease were observed after the second dose^10^. Neutralisating geometric mean titre (GMT) was below cut-off in most cases after prime dose, but as anticipated, titres substantially increased after boost immunization^18^. In older adults mean GMT was only 12 in a preliminary analysis of 12 participants^19^ and increased to 109 after the second dose.

In this study, we assess antibody responses against the the B.1.1.7 variant after vaccination with the first and second doses of BNT162b2, showing modest reduction in neutralisation against pseudoviruses bearing B.1.1.7 Spike mutations (ΔH69/V70, Δ144, N501Y, A570D, P681H, T716I, S982A and D1118H). In addition, by using a panel of human neutralising monoclonal antibodies (mAbs) we show that the B.1.1.7 variant can escape neutralisation mediated by most NTD-specific antibodies tested and by a fraction of RBM-specific antibodies. Finally, we show that the recent emergence and transmission of B.1.1.7 viruses bearing the Spike E484K mutation results in significant additional loss of neutralisation by BNT162b2 mRNA-elicited antibodies, convalescent sera and mAbs.

## Results

Thirty seven participants had received the first dose of BNT162b2 mRNA vaccine three weeks prior to blood draw for serum and peripheral blood monocnulear cells (PBMC) collection. Median age was 63.5 years (IQR 47-84) and 33% were female. Serum IgG titres to Nucleocapsid (N) protein, S and the S receptor binding domain (RBD) were assayed by particle based flow cytometry on a Luminex analyser (**Extended Data Fig. 1a**). These data showed S and RBD antibody titres much higher than in healthy controls, but lower than in individuals recovered from COVID-19 and titres observed in therapeutic convalescent plasma. The raised N titres relative to control could be the result of non-specific cross reactivity that is increased following vaccination. However, the antibody response was heterogeneous with almost 100-fold variation in IgG titres to S and RBD across the vaccinated participants.

Using lentiviral pseudotyping we studied WT (wild type bearing D614G) and mutant B.1.1.7 S proteins (**Fig. 1a**) on the surface of enveloped virions in order to measure neutralisation activity of vaccine-elicited sera. This system has been shown to give results correlating with replication competent authentic virus^20,21^. Eight out of 37 participants exhibited no appreciable neutralisation against the WT pseudotyped virus following the first dose of vaccines. The vaccine sera exhibited a range of inhibitory dilutions giving 50% neutralisation (ID50) (**Fig. 1c-d**). The GMT against wild type (WT) following the second dose of vaccine was an order of magnitude higher than after the first dose (318 vs 77) (Fig 1c-f).There was correlation between full length S IgG titres and serum neutralisation titres (**Extended Data Fig. 1b**). A broad range of T cell responses was measured by IFN gamma FluoroSpot against SARS-CoV-2 peptides in vaccinees. These cellular responses did not correlate with IgG S antibody titres (**Extended Data Fig. 1c-d**).

**Figure 1.**
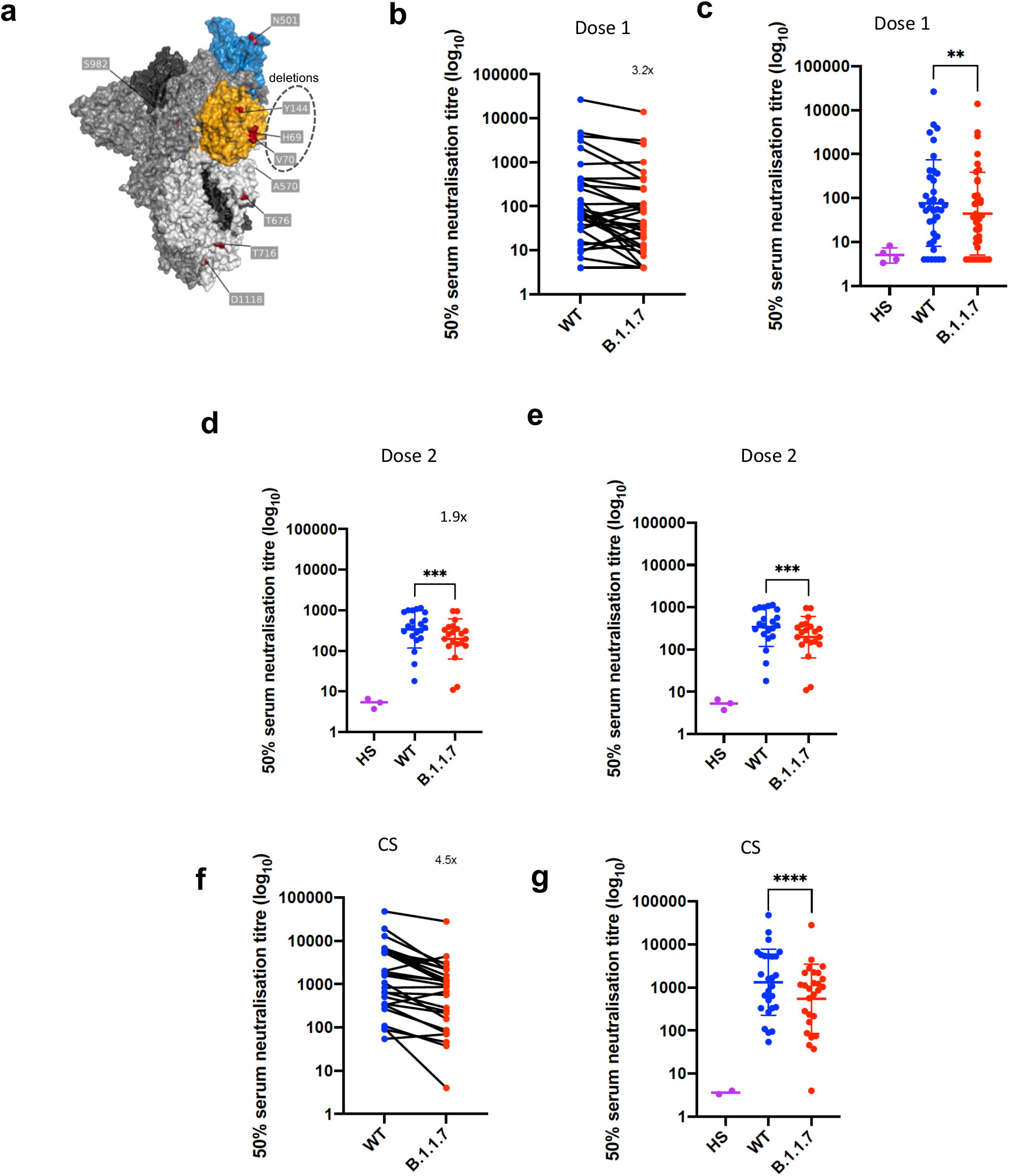
Neutralization by first and second dose mRNA vaccine sera against wild type and B.1.1.7 Spike mutant SARS-CoV-2 pseudotyped viruses. **a**, Spike in open conformation with a single erect RBD (PDB: 6ZGG) in trimer axis vertical view with the locations of mutated residues highlighted in red spheres and labelled on the monomer with erect RBD. Vaccine first dose (**b-c, n=37**), second dose (**d-e, n=21)** and convalescent sera, Conv. (**f-g**,**n=27**) against WT and B.1.1.7 Spike mutant with N501Y, A570D, ΔH69/V70, Δ144/145, P681H, T716I, S982A and D1118H. GMT with s.d presented of two independent experiments each with two technical repeats. Wilcoxon matched-pairs signed rank test p-values * <0.05, ** <0.01, ***<0.001, **** <0.0001, ns not significant HS – human AB serum control. Limit of detection for 50% neutralization set at 10.

We then generated mutated pseudoviruses carrying S protein with mutations N501Y, A570D and the H69/V70 deletion. We observed no reduction in the ability of sera from vaccinees to inhibit either WT or mutant virus (**Extended Data Fig. 2a, b**). A panel of sera from ten recovered individuals also neutralised both wild type and the mutated viruses similarly (**Extended Data Fig. 2c**). We next completed the full set of eight mutations in the S protein present in B.1.1.7 variant (**Fig. 1a**), ΔH69/V70, Δ144, N501Y and A570D in the S_1_ subunit and P681H, T716I, S982A and D1118H in the S_2_ subunit. All constructs also contained D614G. We found that among 29 individuals with neutralisation activity against the WT three weeks after receiving a single dose of the the BNT162b2 mRNA vaccine, 20 showed evidence of reduction in efficacy of antibodies against the B.1.1.7 mutant (**Fig. 1b-c, Extended Data Fig. 3**). The mean fold change reduction in sensitivity to first dose vaccine sera of B.1.1.7 compared to WT was approximately 3.2 (SD 5.7). The variation is likely due to the low neutralisation titres following first dose. Following the second dose, GMT was markedly increased compared with first dose titres, and the mean fold change had reduced to 1.9 (SD 0.9) (**Fig. 1d-e**). Amongst sera from 27 recovered individuals, the GMT at 50% neutralisation was 1334 for WT, significantly higher than post second dose vaccination **(Fig. 1f-g)**. The fold change in ID50 for neutralisation of B.1.1.7 versus wild type (D614G) was 4.5 (**Fig. 1f-g** and **Extended Data Fig. 4**).

### B.1.1.7 with spike E484K mutation and neutralization by vaccine and convalescent sera

The E484K substitution (**Fig. 2a**) is antigenically important, and has been reported as an escape mutation for several monoclonal antibodies including C121, C144, REGN10933 and Ly-CoV555 ^22^. E484K is also known to be present in the B.1.351 (501Y.V2) and P.1 (501Y.V3) lineages in combination with amino acid replacements at N501 and K417. As of 10^th^ Feb 2021, twenty three English and two Welsh B.1.1.7 sequences from viral isolates contained the E484K substitution (**Fig. 2b**). The number of B.1.1.7 sequences has been increasing since the start of December 2020 (**Fig. 2c**). Phylogenetic analysis suggests that there have been multiple independent acquisitions, with one lineage appearing to expand over time, indicating active transmission (**Fig. 2b**). This has resulted in Public Health England naming this as a variant of concern (VOC 202102/02)^23^, triggering enhanced public health measures. There are as yet no phenotypic data on the sensitivity to neutralisation for this virus or its spike protein.

**Figure 2.**
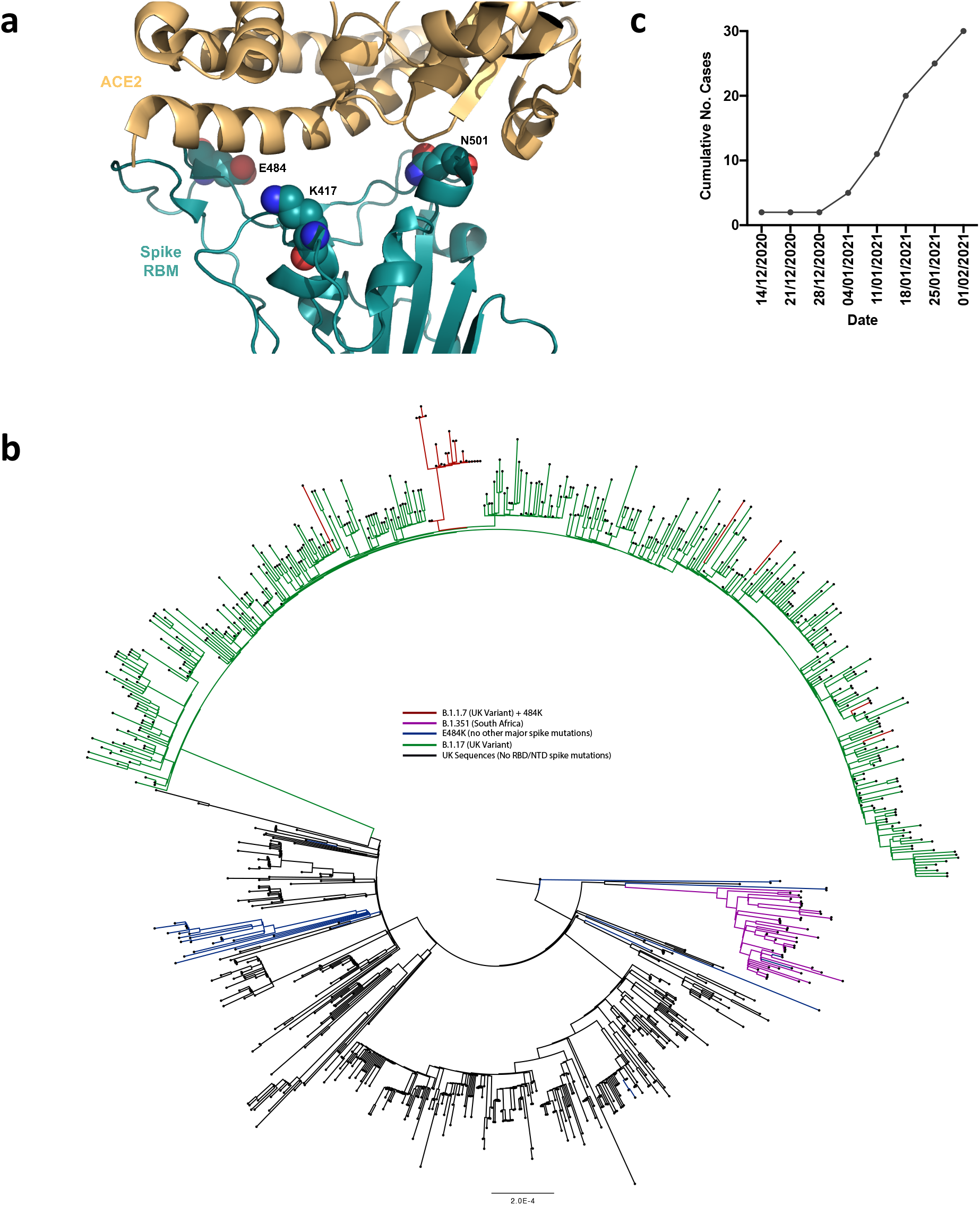
E484K appearing in background of B.1.1.7 with evidence of transmission. **a.** Representation of Spike RBM:ACE2 interface (PDB: 6M0J) with residues E484, N501 and K417 highlighted as spheres coloured by element **b**. Maximum likelihood phylogeny of a subset of sequences from the United Kingdom bearing the E484K mutation (green) and lineage B.1.1.7 (blue), with background sequences without RBD mutations in black. As of 11th Feb 2021, 30 sequences from the B.1.1.7 lineage (one cluster of 25 at top of phylogenetic tree) have acquired the E484K mutation (red). c. Sequence accumulation over time in GISAID for UK sequences with B.1.1.7 and E484K. RBD – receptor binding domain; NTD – N terminal domain.

We therefore generated pseudoviruses bearing B.1.1.7 spike mutations with or without additional E484K and tested these against sera obtained after first and second dose mRNA vaccine as well as against convalescent sera. Following second dose, we observed a significant loss of neutralising activity for the pseudovirus with B.1.1.7 spike mutations and E484K (Fig 3d-e). The mean fold change for the E484K B.1.1.7 Spike was 6.7 compared to 1.9 for B.1.1.7, relative to WT (**Fig. 3a-c**). Similarly when we tested a panel of convalescent sera with a range of neutralisation titres (Fig. 1f-g), we observed additional loss of activity against the mutant B.1.1.7 spike with E484K, with fold change of 11.4 relative to WT (**Fig. 3f-g)**.

**Figure 3.**
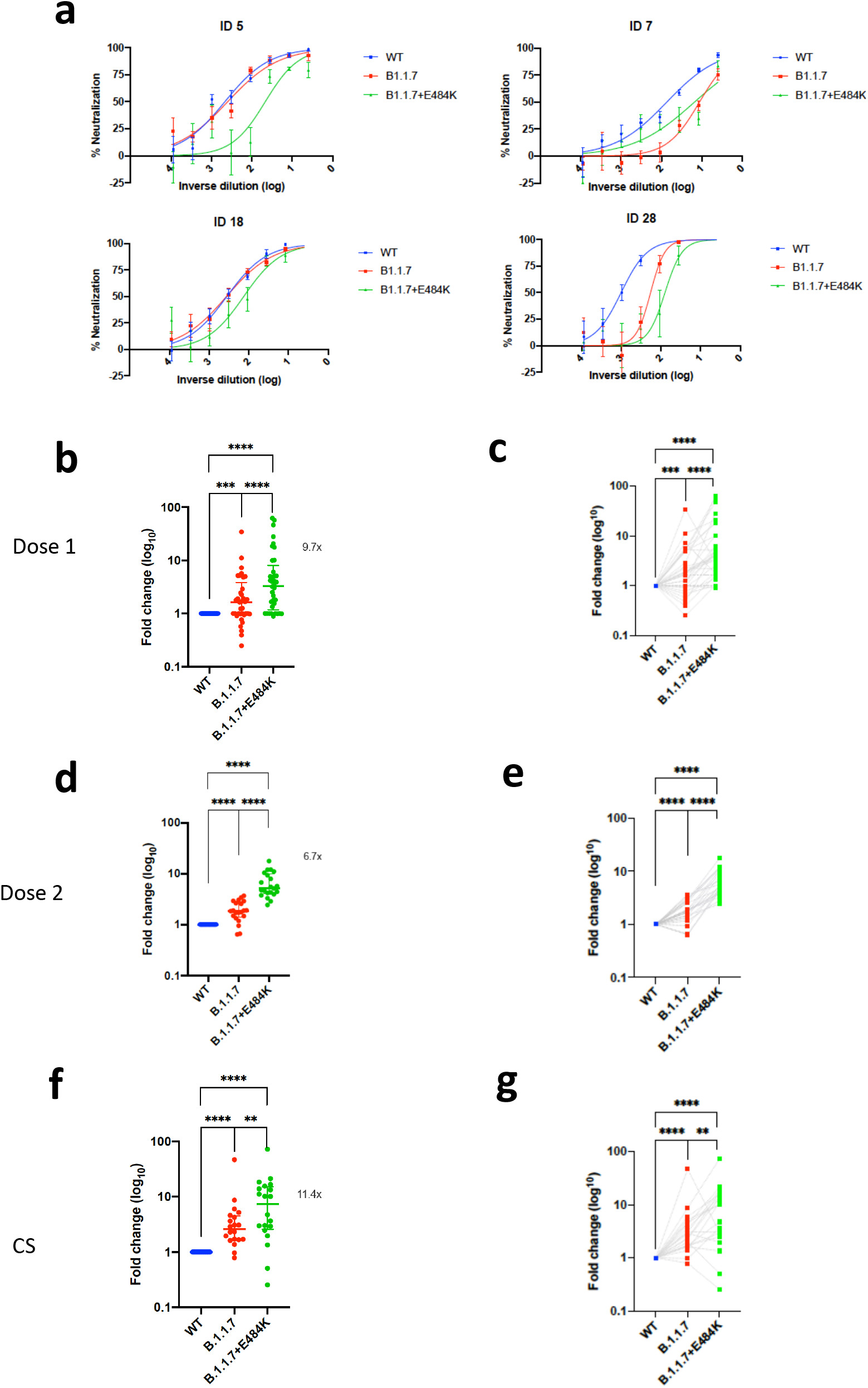
Neutralization potency of mRNA vaccine sera and convalescent sera (pre SARS-CoV-2 B.1.1.7) against pseudotyped virus bearing Spike mutations in the B1.1.7 lineage with and without E484K in the receptor binding domain (all In Spike D614G background). **a**, Example neutralization curves for vaccinated individuals. Data points represent mean of technical replicates with standard error and are representative of two independent experiments (**b-g)**. 50% neutralisation titre for each virus against sera derived (b,c, n=37) following first vaccination (d,e, n=21) following second vaccination and (f,g, n=20) convalescent sera (CS) expressed as fold change relative to WT. Data points are mean fold change of technical replicates and are representative of two independent experiments. Central bar represents mean with outer bars representing s.d. Wilcoxon matched-pairs signed rank test p-values *<0.05, **<0.01, ***<0.001, ****<0.0001; ns not significant. Limit of detection for 50% neutralization set at 10.

### B.1.1.7 variant escape from NTD- and RBM-specific mAb-mediated neutralization

To investigate the role of the full set of mutations in NTD, RBD and S2 present in the B.1.1.7 variant, we tested 60 mAbs isolated from 15 individuals that recovered from SARS-CoV-2 infection in early 2020 with an *in-vitro* pseudotyped neutralization assay using VeroE6 target cells expressing Transmembrane protease serine 2 (TMPRSS2, **Extended Data Table 1**). We found that 20 out of 60 (33.3%) mAbs showed a greater than 2-fold loss of neutralising activity of B.1.1.7 variant compared to WT SARS-CoV-2 (**Fig. 4a,b** and **Extended Data Fig. 5**). Remarkably, the B.1.1.7 mutant virus was found to fully escape neutralization by 8 out of 10 NTD-targeting mAbs (80%), and partial escape from an additional mAb (10%) (**Fig. 4c**). We previously showed that the deletion of residue 144 abrogates binding by 4 out of 6 NTD-specific mAbs tested, possibly accounting for viral neutralization escape by most NTD-specific antibodies^24^. Of the 31 RBM-targeting mAbs, 5 (16.1%) showed more than 100-fold decrease in B.1.1.7 neutralization, and additional 6 mAbs (19.4%) had a partial 2-to-10-fold reduction (**Fig. 4d**). Finally, all RBD-specific non-RBM-targeting mAbs tested fully retained B.1.1.7 neutralising activity (**Fig. 4e**).

**Figure 4.**
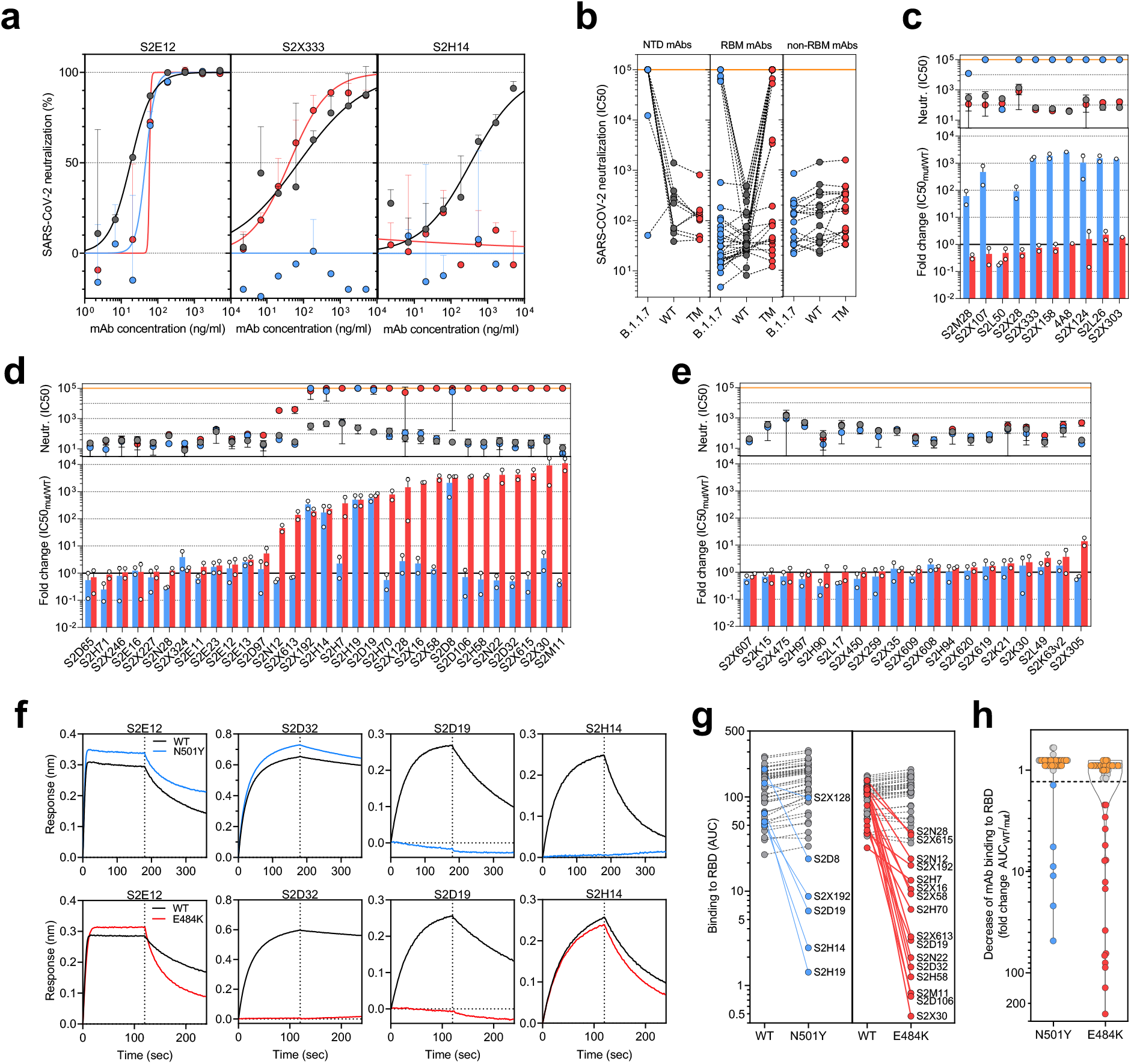
Neutralization and binding by a panel of NTD- and RBD-specific mAbs against WT, B.1.1.7 and RBD mutant SARS-CoV-2 viruses. **a**, Neutralization of WT D614G (black), B.1.1.7 (blue) and a triple mutant (TM, carrying RBD mutations K417N/E484K/N501Y) (red) pseudotyped SARS-CoV-2-MLVs by 3 selected mAbs (S2E12, S2×333 and S2H14) from one representative experiment. Shown is the mean ± s.d. of 2 technical replicates. **b**, Neutralization of WT (D614G), B.1.1.7 and TM SARS-CoV-2-MLVs by 60 mAbs targeting NTD (n=10), RBM (n=31) and non-RBM sites in the RBD (n=19). Shown are the mean IC50 values (ng/ml) of n=2 independent experiments. **c-e**, Neutralization shown as mean IC50 values (upper panel) and mean fold change of B.1.1.7 (blue) or TM (red) relative to WT (lower panel) of NTD (c), RBM (d) and non-RBM (e) mAbs. Lower panel shows IC50 values from 2 independent experiments. **f-h**, Kinetics of binding of mAbs to WT (black), N501Y (blue) and E484K (red) RBD as measured by bio-layer interferometry (BLI). Shown in (f) are the 4 RBM-targeting mAbs with no reduced binding to N501Y or E484K RBD. Area under the curve (AUC) (g) and AUC fold change (h) of 50 mAbs tested against WT, N501Y and E484K RBD. mAbs with a >1.3 AUC fold change shown in blue and red. mAbs: monoclonal antibodies. NTD: N- terminal domain

To address the role of B.1.1.7 N501Y mutation in the neutralization escape from RBM-specific antibodies, we tested the binding of 50 RBD-specific mAbs to WT and N501Y mutant RBD by biolayer interferometry (**Fig. 4f** and **Extended Data Fig. 6**). The 5 RBM-specific mAbs that failed to neutralize B.1.1.7 variant (**Fig. 4d**) showed a complete loss of binding to N501Y RBD mutant (**Fig. 4g-h**), demonstrating a role for this mutation as an escape mechanism for certain RBM-targeting mAbs.

The decreased neutralising activity of the immune sera from vaccinees and convalescent patients against B.1.1.7, but not against Δ69/70-501Y-570D mutant (**Fig. 1** and **Extended Data Fig. 2**), could be the result of a loss of neutralising activity of both RBD- and NTD-targeting antibodies, and suggests that the key mutation is Δ144. RBD antibodies against N501Y could play a role in decreased neutralisation by sera, with the overall impact possibly modulated by other mutations present in B.1.1.7, as well as the relative dominance of NTD versus RBM antibodies in polyclonal sera.

To assess the effect of E484K on this panel of mAbs we generated a SARS-CoV-2 pseudotype carrying the K417N, E484K and N501Y mutations (TM). The inclusion of the K417N substitution was prompted by the observation that substitutions at this position have been found in 5 sequences from recent viral isolates within the B.1.1.7 lineage (K417 to N, E or R). This is in keeping with convergent evolution of the virus towards an RBD with N501Y, E484K and K417N/T as evidenced by B.1.351 and P.1 lineages (K417N or K417T, respectively) causing great concern globally. It is therefore important to assess this combination going forward.

Importantly, mutations at K417 are reported to escape neutralization from mAbs, including the recently approved mAb LY-CoV016 ^22,25^. Out of the 60 mAbs tested, 20 (33.3%) showed >10 fold loss of neutralising activity of TM mutant compared to WT SARS-CoV-2 (**Fig. 4 a-b** and **Extended Data Fig. 5**), and of these 19 are RBM-specific mAbs. As above, we addressed the role of E484K mutation in escape from RBM-specific antibodies, by testing the binding of 50 RBD-specific mAbs to WT and E484K mutant RBD by biolayer interferometry (**Fig. 4f** and **Extended Data Fig. 7**). Out of the 19 RBM-specific mAbs that showed reduced or loss of neutralization of TM mutant (**Fig. 4d**), 16 showed a complete or partial loss of binding to E484K RBD mutant (**Fig. 4g-h**), consistent with findings that E484K is an important viral escape mutation^26, 39, 27^. Three of these 16 mAbs also lost binding to an RBD carrying N501Y, indicating that a fraction of RBM antibodies are sensitive to both N501Y and E484K mutations. Similarly, 3 of the 19 mAbs that lost neutralization of TM mutant (S2D8, S2H7 and S2×128) were previously shown to lose binding and neutralization to the K417V mutant, and here shown to be sensitive to either N501Y or E484K mutations.

### SARS-CoV-2 B.1.1.7 binds human ACE2 with higher affinity than WT

SARS-CoV-2 and SARS-CoV enter host cells through binding of the S glycoprotein to angiotensin converting enzyme 2 (ACE2)^1,28^. Previous studies showed that the binding affinity of SARS-CoV for human ACE2 correlated with the rate of viral replication in distinct species, transmissibility and disease severity ^29-31^. However, the picure is unclear for SARS-CoV-2. To understand the potential contribution of receptor interaction to infectivity, we set out to evaluate the influence of the B.1.1.7 RBD substitution N501Y on receptor engagement. We used biolayer interferometry to study binding kinetics and affinity of the purified human ACE2 ectodomain (residues 1-615) to immobilized biotinylated SARS-CoV-2 B.1.1.7 or WT RBDs. We found that ACE2 bound to the B.1.1.7 RBD with an affinity of 22 nM compared to 133 nM for the WT RBD **(Extended Data Fig. 8)**, in agreement with our previous deep-mutational scanning measurements using dimeric ACE2^32^. Although ACE2 bound with comparable on-rates to both RBDs, the observed dissociation rate constant was slower for B.1.1.7 than for the WT RBD **(Table 1)**.

**Table 1.** **Kinetic analysis of human ACE2 binding to SARS-CoV-2 Wuhan-1, N501Y and N501Y/ E484K/ K417N (TM) RBDs by biolayer interferometry**. Values reported represent the global fit to the data shown in Extended Data Fig. 8.

|  |  | SARS-CoV-2 RBD WT | SARS-CoV-2 RBD N501Y | SARS-CoV-2 RBD TM |
| --- | --- | --- | --- | --- |
| **K<sub>D</sub> (nM)** | hACE2 | 133 | 22 | 64 |
| **k<sub>on</sub> (M<sup>-1</sup>.s<sup>-1</sup>)** |  | 1.3*10<sup>5</sup> | 1.4*10<sup>5</sup> | 1.3*10<sup>5</sup> |
| **k<sub>off</sub> (s<sup>-1</sup>)** |  | 1.8*10<sup>-2</sup> | 3*10<sup>-3</sup> | 8.5*10<sup>-3</sup> |

To understand the impact of TM mutations (K417N, E484K and N501Y), we evaluated binding of ACE2 to the immobilized TM RBD using biolayer interferometry. We determined an ACE2 binding affinity of 64 nM for the TM RBD which is driven by a faster off-rate than observed for the B.1.1.7 RBD but slower than for the WT RBD. Based on our previous deep-mutational scanning measurements using dimeric ACE2, we propose that the K417N mutation is slightly detrimental to ACE2 binding explaining the intermediate affinity determined for the TM RBD compared to the B.1.17 and WT RBDs, likely as a result of disrupting the salt bridge formed with ACE2 residue D30. Enhanced binding of the B.1.1.7 RBD to human ACE2 resulting from the N501Y mutation might participate in the efficient ongoing transmission of this newly emergent SARS-CoV-2 lineage, and possibly reduced opportunity for antibody binding. Although the TM RBD mutations found in B.1.351 are known to participate in immune evasion^33,34^, the possible contribution to transmissibility of enhanced ACE2 binding relative to WT remains to be determined for this lineage.

## Discussion

Serum neutralising activity is a correlate of protection for other respiratory viruses, including influenza^35^ and respiratory syncytial virus where prohylaxis with monoclonal antibodies has been used in at-risk groups^36,37^. Neutralising antibody titres appeared to be highly correlated with vaccine protection against SARS-CoV-2 rechallenge in non-human primates, and importantly, there was no correlation between T cell responses (as measured by ELISpot) and protection^38^. Moreover, passive transfer of purified polyclonal IgGs from convalescent macaques protected naïve macaques against subsequent SARS-CoV-2 challenge^39^. Coupled with multiple reports of re-infection, there has therefore been significant attention placed on virus neutralisation.

This study reports on the neutralisation by sera collected after both the first and second doses of the BNT162b2 vaccine. The participants of this study were older adults, in line with the targeting of this age group in the initial rollout of the vaccination campaign in the UK. Participants showed similar neutralising activity against wild type pseudovirus as in the phase I/II study^12^. This is relevant for the UK and other countries planning to extend the gap between doses of mRNA and adenovirus based vaccines from 3 to 12 weeks, despite lack of data for this schedule for mRNA vaccines in particular.

The three mutations in S1 (N501Y, A570D, ΔH69/V70) did not appear to impact neutralisation in a pseudovirus assay, consistent with data on N501Y having little effect on nuetralisation by convalescent and post vaccination sera^40^. However, we demonstrated that a pseudovirus bearing S protein with the full set of mutations present in the B.1.1.7 variant (i.e., ΔH69/V70, Δ144, N501Y, A570D, P681H, T716I, S982A, D1118H) did result in small reduction in neutralisation by sera from vaccinees that was more marked following the first dose than the second dose. This could be related to increased breadth/potency/concentration of antibodies following the boost dose. A reduction in neutralization titres from mRNA-elicited antibodies in volunteers who received two doses (using both mRNA-1273 and BNT162b2 vaccines) was also observed by Wang et al.^41^ using pseudoviruses carrying the N501Y mutation. Other studies also reported small reduction of neutralization against the B.1.1.7 variant against sera from individuals vaccinated with two doses of BNT162b2^42^ and mRNA-1273^43^. Xie et al did not find an effect of N501Y alone in the context of BNT162b2 vaccine sera^44^.

The reduced neutralising activity observed with polyclonal antibodies elicited by mRNA vaccines observed in this study is further supported by the loss of neutralising activity observed with human mAbs directed to both RBD and, to a major extent, to NTD. In the study by Wang et al., 6 out 17 RDB-specific mAbs isolated from mRNA-1273 vaccinated individuals showed more than 100-fold neutralisation loss against N501Y mutant, a finding that is consistent with the loss of neutralisation by 5 out 29 RBM-specific mAbs described in this study. However, the contribution of N501Y to loss of neutralisation activity of polyclonal vaccine and convalescent sera is less clear, and interactions with other mutations likely.

Multiple variants, including the 501Y.V2 and B.1.1.7 lineages, harbor multiple mutations as well as deletions in NTD, most of which are located in a site of vulnerability that is targeted by all known NTD-specific neutralising antibodies^24,45^. The role of NTD-specific neutralising antibodies might be under-estimated, in part by the use of neutralization assays based on target cells over-expressing ACE2 receptor. NTD-specific mAbs were suggested to interfere with viral entry based on other accessory receptors, such as DC-SIGN and L-SIGN^46^, and their neutralization potency was found to be dependent on different in vitro culture conditions^24^. The observation that 9 out of 10 NTD-specific neutralising antibodies failed to show a complete or near-complete loss of neutralising activity against B.1.1.7 indicates that this new variant may have evolved also to escape from this class of antibodies, that may have a yet unrecognized role in protective immunity. Wibmer et al.^34^ have also recently reported the loss of neutralization of 501Y.V2 by the NTD-specific mAb 4A8, likely driven by the R246I mutation. This result is in line with the lack of neutralization of B.1.1.7 by the 4A8 mAb observed in this study, likely caused by Δ144 due to loss of binding^24^. Finally, the role of NTD mutations (in particular, L18F, Δ242-244 and R246I) was further supported by the marked loss of neutralization observed by Wibmer et al.^34^ against 501Y.V2 compared to the chimeric pseudotyped viral particle carrying only the RBD mutations K417N, E484K and N501Y. Taken together, the presence of multiple escape mutations in NTD is supportive of the hypothesis that this region of the spike, in addition to RBM, is also under immune pressure.

Worryingly, we have shown that there are multiple B.1.1.7 sequences in the UK bearing E484K with early evidence of transmission as well as independent aquisitions. We measured further reduction neutralisation titers by vaccine sera when E484K was present alongside the B.1.1.7 S mutations. Wu and co-authors^43^ have also shown that variants carrying the E484K mutation resulted in 3-to-6 fold reduction in neutralization by sera from mRNA-1273 vaccinated individuals. Consistently, in this study we found that approximately 50% of the RBM mAbs tested lost neutralising activity against SARS-CoV-2 carrying E484K. E484K has been shown to impact neutralisation by monoclonal antibodies or convalescent sera, especially in combination with N501Y and K417N^16,26,47-49^. Wang et al also showed reduced neutralisation by mRNA vaccine sera against E484K bearing pseudovirus^34^.

Evidence for the importance role of NTD deletions in combination with E484K in immune escape is provided by Andreano *et al*.^27^ who describe the emergence of Δ140 in virus co-incubated with potently neutralising convalescent plasma, causing a 4-fold reduction in neutralization titre. This Δ140 mutant subsequently acquired E484K which resulted in a further 4-fold drop in neutralization titre indicating a two residue change across NTD and RBD represents an effective pathway of escape that can dramatically inhibit the polyclonal response.

Our study was limited by modest sample size. Although the spike pseudotyping system has been shown to faithfully represent full length infectious virus, there may be determinants outside the S that influence escape from antibody neutralization either directly or indirectly in a live replication competent system. On the other hand live virus systems allow replication and therefore mutations to occur, and rigorous sequencing at multiple steps is needed.

Vaccines are a key part of a long term strategy to bring SARS-CoV-2 transmission under control. Our data suggest that vaccine escape to current Spike directed vaccines designed against the Wuhan strain will be inevitable, particularly given that E484K is emerging independently and recurrently on a B.1.1.7 (501Y.V1) background, and given the rapid global spread of B.1.1.7. Other major variants with E484K such as 501Y.V2 and V3 are also spreading regionally. This should be mitigated by designing next generation vaccines with mutated S sequences and using alternative viral antigens.

## Data Availability

Data are available from the corresponding author on request

## Acknowledgements

We would like to thank Cambridge University Hospitals NHS Trust Occupational Health Department. We would also like to thank the NIHR Cambridge Clinical Research Facility and staff at CUH and. We would like to thank Eleanor Lim and Georgina Okecha. We thank Dr James Voss for the kind gift of HeLa cells stably expressing ACE2. RKG is supported by a Wellcome Trust Senior Fellowship in Clinical Science (WT108082AIA). LEM is supported by a Medical Research Council Career Development Award (MR/R008698/1). SAK is supported by the Bill and Melinda Gates Foundation via PANGEA grant: OPP1175094. DAC is supported by a Wellcome Trust Clinical PhD Research Fellowship. KGCS is the recipient of a Wellcome Investigator Award (200871/Z/16/Z). This research was supported by the National Institute for Health Research (NIHR) Cambridge Biomedical Research Centre, the Cambridge Clinical Trials Unit (CCTU), and the NIHR BioResource. This study was supported by the National Institute of General Medical Sciences (R01GM120553 to D.V.), the National Institute of Allergy and Infectious Diseases (DP1AI158186 and HHSN272201700059C to D.V.), a Pew Biomedical Scholars Award (D.V.), an Investigators in the Pathogenesis of Infectious Disease Awards from the Burroughs Wellcome Fund (D.V.) and Fast Grants (D.V.). The views expressed are those of the authors and not necessarily those of the NIHR or the Department of Health and Social Care. JAGB is supported by the Medical Research Council (MC_UP_1201/16). IATM is funded by a SANTHE award.

## Author contributions

Conceived study: D.C., RKG, DAC. Designed study and experiments: RKG, DAC, LEM, JB, MW, JT, LCG, GBM, RD, BG, NK, AE, M.P., D.V., L.P., A.D.M, J.B., D.C. Performed experiments: BM, DAC, RD, IATMF, ACW, LCG, GBM. Interpreted data: RKG, DAC, BM, RD, IATMF, ACW, LEM, JB, KGCS, DV. ADM, JB and CSF carried out pseudovirus neutralization assays. DP produced pseudoviruses. MSP, LP, DV and DC designed the experiments. MAT, JB, NS and SJ expressed and purified the proteins. KC, SJ and EC sequenced and expressed antibodies. EC and KC performed mutagenesis for mutant expression plasmids. ACW and S.B. performed binding assays. AR, AFP and CG contributed to donor’s recruitment and sample collection related to mAbs isolation. HWV, GS, AL, DV, LP, DV and DC analyzed the data and prepared the manuscript with input from all authors.

## MATERIALS AND METHODS

### Participant recruitment and ethics

Participants who had received the first dose of vaccine and individuals with COVID-19 (Coronavirus Disease-19) were consented into the COVID-19 cohort of the NIHR Bioresource. The study was approved by the East of England – Cambridge Central Research Ethics Committee (17/EE/0025).

### SARS-CoV-2 serology by multiplex particle-based flow cytometry (Luminex)

Recombinant SARS-CoV-2 N, S and RBD were covalently coupled to distinct carboxylated bead sets (Luminex; Netherlands) to form a 3-plex and analyzed as previously described (Xiong et al. 2020). Specific binding was reported as mean fluorescence intensities (MFI). Linear regression was used to explore the association between antibody response, T cell response and serum neutralisation in Stata 13. The Pearson correlation coefficient was reported.

### Recombinant expression of SARS-CoV-2-specific mAbs

Human mAbs were isolated from plasma cells or memory B cells of SARS-CoV-2 immune donors, as previously described ^50-52^. Recombinant antibodies were expressed in ExpiCHO cells at 37°C and 8% CO_2_. Cells were transfected using ExpiFectamine. Transfected cells were supplemented 1 day after transfection with ExpiCHO Feed and ExpiFectamine CHO Enhancer. Cell culture supernatant was collected eight days after transfection and filtered through a 0.2 µm filter. Recombinant antibodies were affinity purified on an ÄKTA xpress fast protein liquid chromatography (FPLC) device using 5 mL HiTrap™ MabSelect™ PrismA columns followed by buffer exchange to Histidine buffer (20 mM Histidine, 8% sucrose, pH 6) using HiPrep 26/10 desalting columns

## Generation of S mutants

Amino acid substitutions were introduced into the D614G pCDNA_SARS-CoV-2_S plasmid as previously described^53^ using the QuikChange Lightening Site-Directed Mutagenesis kit, following the manufacturer’s instructions (Agilent Technologies, Inc., Santa Clara, CA). Sequences were checked by Sanger sequencing.

Preparation of B.1.1.7 or TM SARS-CoV-2 S glycoprotein-encoding-plasmid used to produce SARS-CoV-2-MLV based on overlap extension PCR. Briefly, a modification of the overlap extension PCR protocol^54^ was used to introduce the nine mutations of the B.1.1.7 lineage or the three mutations in TM mutant in the SARS-CoV-2 S gene. In a first step, 9 DNA fragments with overlap sequences were amplified by PCR from a plasmid (phCMV1, Genlantis) encoding the full-length SARS-CoV-2 S gene (BetaCoV/Wuhan-Hu-1/2019, accession number mn908947). The mutations (del-69/70, del-144, N501Y, A570D, D614G, P681H, S982A, T716I and D1118H or K417N, E484K and N501Y) were introduced by amplification with primers with similar Tm. Deletion of the C-terminal 21 amino acids was introduced to increase surface expression of the recombinant S^55^. Next, 3 contiguous overlapping fragments were fused by a first overlap PCR (step 2) using the utmost external primers of each set, resulting in 3 larger fragments with overlapping sequences. A final overlap PCR (step 3) was performed on the 3 large fragments using the utmost external primers to amplify the full-length S gene and the flanking sequences including the restriction sites KpnI and NotI. This fragment was digested and cloned into the expression plasmid phCMV1. For all PCR reactions the Q5 Hot Start High fidelity DNA polymerase was used (New England Biolabs Inc.), according to the manufacturer’s instructions and adapting the elongation time to the size of the amplicon. After each PCR step the amplified regions were separated on agarose gel and purified using Illustra GFX™ PCR DNA and Gel Band Purification Kit (Merck KGaA).

### Pseudotype virus preparation

Viral vectors were prepared by transfection of 293T cells by using Fugene HD transfection reagent (Promega). 293T cells were transfected with a mixture of 11ul of Fugene HD, 1µg of pCDNAΔ19spike-HA, 1ug of p8.91 HIV-1 gag-pol expression vector^56,57^, and 1.5µg of pCSFLW (expressing the firefly luciferase reporter gene with the HIV-1 packaging signal). Viral supernatant was collected at 48 and 72h after transfection, filtered through 0.45um filter and stored at −80°C. The 50% tissue culture infectious dose (TCID50) of SARS-CoV-2 pseudovirus was determined using Steady-Glo Luciferase assay system (Promega).

### Serum/plasma pseudotype neutralization assay

Spike pseudotype assays have been shown to have similar characteristics as neutralisation testing using fully infectious wild type SARS-CoV-2^20^. Virus neutralisation assays were performed on 293T cell transiently transfected with ACE2 and TMPRSS2 using SARS-CoV-2 spike pseudotyped virus expressing luciferase^58^. Pseudotyped virus was incubated with serial dilution of heat inactivated *human s*erum samples or sera from vaccinees in duplicate for 1h at 37°C. Virus and cell only controls were also included. Then, freshly trypsinized 293T ACE2/TMPRSS2 expressing cells were added to each well. Following 48h incubation in a 5% CO2 environment at 37°C, luminescence was measured using the Steady-Glo or Bright-Glo Luciferase assay system (Promega). Neutralization was calculated relative to virus only controls. Dilution curves were presented as a mean neutralization with standard error of the mean (SEM). ID50 values were calculated in GraphPad Prism. The ID50 withing groups were summarised as a geometric mean titre and statistical comparison between groups were made with Wilxocon ranked sign test. In addition, the impact of the mutations on the neutralising effect of the sera were expressed as fold change (FC) of ID50 of the wild-type compared to mutant pseudotyped virus. Statistical difference in the mean FC between groups was determined using a 2-tailed t-test.

### IFNγ FluoroSpot assays

Frozen PBMCs were rapidly thawed, and the freezing medium was diluted into 10ml of TexMACS media (Miltenyi Biotech), centrifuged and resuspended in 10ml of fresh media with 10U/ml DNase (Benzonase, Merck-Millipore via Sigma-Aldrich), PBMCs were incubated at 37°C for 1h, followed by centrifugation and resuspension in fresh media supplemented with 5% Human AB serum (Sigma Aldrich) before being counted. PBMCs were stained with 2ul of each antibody: anti-CD3-fluorescein isothiocyanate (FITC), clone UCHT1; anti-CD4-phycoerythrin (PE), clone RPA-T4; anti-CD8a-peridinin-chlorophyll protein - cyanine 5.5 (PerCP Cy5.5), clone RPA-8a (all BioLegend, London, UK), LIVE/DEAD Fixable Far Red Dead Cell Stain Kit (Thermo Fisher Scientific). PBMC phenotyping was performed on the BD Accuri C6 flow cytometer. Data were analysed with FlowJo v10 (Becton Dickinson, Wokingham, UK). 1.5 to 2.5 × 105 PBMCs were incubated in pre-coated Fluorospot plates (Human IFNγ FLUOROSPOT (Mabtech AB, Nacka Strand, Sweden)) in triplicate with peptide mixes specific for Spike, Nucleocapsid and Membrane proteins of SARS-CoV-2 (final peptide concentration 1µg/ml/peptide, Miltenyi Biotech) and an unstimulated and positive control mix (containing anti-CD3 (Mabtech AB), Staphylococcus Enterotoxin B (SEB), Phytohaemagglutinin (PHA) (all Sigma Aldrich)) at 37°C in a humidified CO2 atmosphere for 48 hours. The cells and medium were decanted from the plate and the assay developed following the manufacturer’s instructions. Developed plates were read using an AID iSpot reader (Oxford Biosystems, Oxford, UK) and counted using AID EliSpot v7 software (Autoimmun Diagnostika GmbH, Strasberg, Germany). All data were then corrected for background cytokine production and expressed as spot forming units (SFU)/Million PBMC or CD3 T cells. The association between spike Tcell response, spike specific antibody response and serum neutralisation was deterimined using linear regression and the Pearson correlation coefficient between these variables were determined using Stata 13.

### Ab discovery and recombinant expression

Human mAbs were isolated from plasma cells or memory B cells of SARS-CoV or SARS-CoV-2 immune donors, as previously described ^48,56-58^. Recombinant antibodies were expressed in ExpiCHO cells at 37°C and 8% CO2. Cells were transfected using ExpiFectamine. Transfected cells were supplemented 1 day after transfection with ExpiCHO Feed and ExpiFectamine CHO Enhancer. Cell culture supernatant was collected eight days after transfection and filtered through a 0.2 µm filter. Recombinant antibodies were affinity purified on an ÄKTA xpress FPLC device using 5 mL HiTrap™ MabSelect™ PrismA columns followed by buffer exchange to Histidine buffer (20 mM Histidine, 8% sucrose, pH 6) using HiPrep 26/10 desalting columns.

### MAbs pseudovirus neutralization assay

MLV-based SARS-CoV-2 S-glycoprotein-pseudotyped viruses were prepared as previously described (Pinto et al., 2020). HEK293T/17cells were cotransfected with a WT, B.1.1.7 or TM SARS-CoV-2 spike glycoprotein-encoding-plasmid, an MLV Gag-Pol packaging construct and the MLV transfer vector encoding a luciferase reporter using X-tremeGENE HP transfection reagent (Roche) according to the manufacturer’s instructions. Cells were cultured for 72 h at 37°C with 5% CO_2_ before harvesting the supernatant. VeroE6 stably expressing human TMPRSS2 were cultured in Dulbecco’s Modified Eagle’s Medium (DMEM) containing 10% fetal bovine serum (FBS), 1% penicillin–streptomycin (100 I.U. penicillin/mL, 100 µg/mL), 8 µg/mL puromycin and plated into 96-well plates for 16–24 h. Pseudovirus with serial dilution of mAbs was incubated for 1 h at 37°C and then added to the wells after washing 2 times with DMEM. After 2–3 h DMEM containing 20% FBS and 2% penicillin–streptomycin was added to the cells. Following 48-72 h of infection, Bio-Glo (Promega) was added to the cells and incubated in the dark for 15 min before reading luminescence with Synergy H1 microplate reader (BioTek). Measurements were done in duplicate and relative luciferase units were converted to percent neutralization and plotted with a non-linear regression model to determine IC50 values using GraphPad PRISM software (version 9.0.0).

### Antibody binding measurements using bio-layer interferometry (BLI)

MAbs were diluted to 3 µg/ml in kinetic buffer (PBS supplemented with 0.01% BSA) and immobilized on Protein A Biosensors (FortéBio). Antibody-coated biosensors were incubated for 3□min with a solution containing 5□µgL□/ml of WT, N501Y or E484K SARS-CoV-2 RBD in kinetic buffer, followed by a 3-min dissociation step. Change in molecules bound to the biosensors caused a shift in the interference pattern that was recorded in real time using an Octet RED96 system (FortéBio). The binding response over time was used to calculate the area under the curve (AUC) using GraphPad PRISM software (version 9.0.0).

### Production of SARS-CoV-2 and B.1.1.7 receptor binding domains and human ACE2

The SARS-CoV-2 RBD (BEI NR-52422) construct was synthesized by GenScript into CMVR with an N-terminal mu-phosphatase signal peptide and a C-terminal octa-histidine tag (GHHHHHHHH) and an avi tag. The boundaries of the construct are N-_328_RFPN_331_ and _528_KKST_531_-C^59^. The B.1.1.7 RBD gene was synthesized by GenScript into pCMVR with the same boundaries and construct details with a mutation at N501Y. These plasmids were transiently transfected into Expi293F cells using Expi293F expression medium (Life Technologies) at 37°C 8% CO_2_ rotating at 150 rpm. The cultures were transfected using PEI cultivated for 5 days. Supernatants were clarified by centrifugation (10 min at 4000xg) prior to loading onto a nickel-NTA column (GE). Purified protein was biotinylated overnight using BirA (Biotin ligase) prior to size exclusion chromatography (SEC) into phosphate buffered saline (PBS). Human ACE2-Fc (residues 1-615 with a C-terminal thrombin cleavage site and human Fc tag) were synthesized by Twist. Clarified supernatants were affinity purified using a Protein A column (GE LifeSciences) directly neutralized and buffer exchanged. The Fc tag was removed by thrombin cleavage in a reaction mixture containing 3 mg of recombinant ACE2-FC ectodomain and 10 μg of thrombin in 20 mM Tris-HCl pH8.0, 150 mM NaCl and 2.5 mM CaCl_2_.The reaction mixture was incubated at 25°C overnight and re-loaded on a Protein A column to remove uncleaved protein and the Fc tag. The cleaved protein was further purified by gel filtration using a Superdex 200 column 10/300 GL (GE Life Sciences) equilibrated in PBS.

### Protein affinity measurement using bio-layer interferometry

Biotinylated RBD (WT, N501Y, or TM) were immobilized at 5 ng/uL in undiluted 10X Kinetics Buffer (Pall) to SA sensors until a load level of 1.1nm. A dilution series of either monomeric ACE2 or Fab in undiluted kinetics buffer starting at 1000-50nM was used for 300-600 seconds to determine protein-protein affinity. The data were baseline subtracted and the plots fitted using the Pall FortéBio/Sartorius analysis software (version 12.0). Data were plotted in Prism.

#### Statistical analysis

Linear regression was used to explore the association between antibody response, T cell response and serum neutralisation in Stata 13. The Pearson correlation coefficient was reported.

#### Neutralisation data analysis

Neutralization was calculated relative to virus only controls. Dilution curves were presented as a mean neutralization with standard error of the mean (SEM). IC50 values were calculated in GraphPad Prism. The inhibitory dilution (ID50) within groups were summarised as a geometric mean titre and statistical comparison between groups were made with Wilxocon ranked sign test. In addition, the impact of the mutations on the neutralising effect of the sera were expressed as fold change of ID50 of the wild-type compared to mutant pseudotyped virus. Statistical difference in the mean FC between groups was determined using a 2-tailed t-test

#### IFNγ FluoroSpot assay data analysis

The association between spike Tcell response, spike specific antibody response and serum neutralisation was determined using linear regression and the Pearson correlation coefficient between these variables were determined using Stata 13.

## Data availability

The neutralization and BLI data shown in Fig. 4 and Extended Data Fig. 5-7 can be found in **Source Data Fig. 4**. Other data are available from the corresponding author on request.

---

**Extended Data Table 1.**
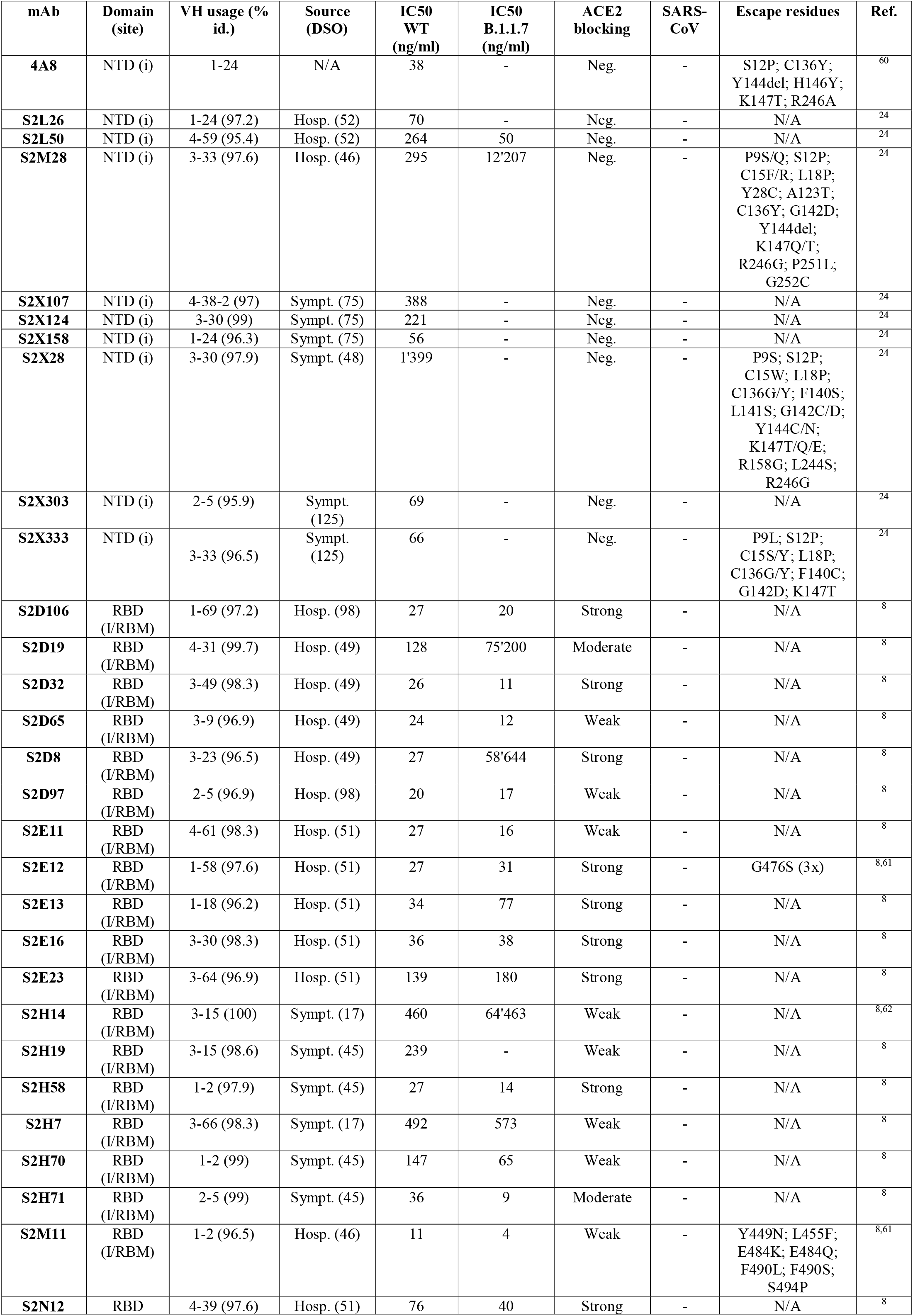
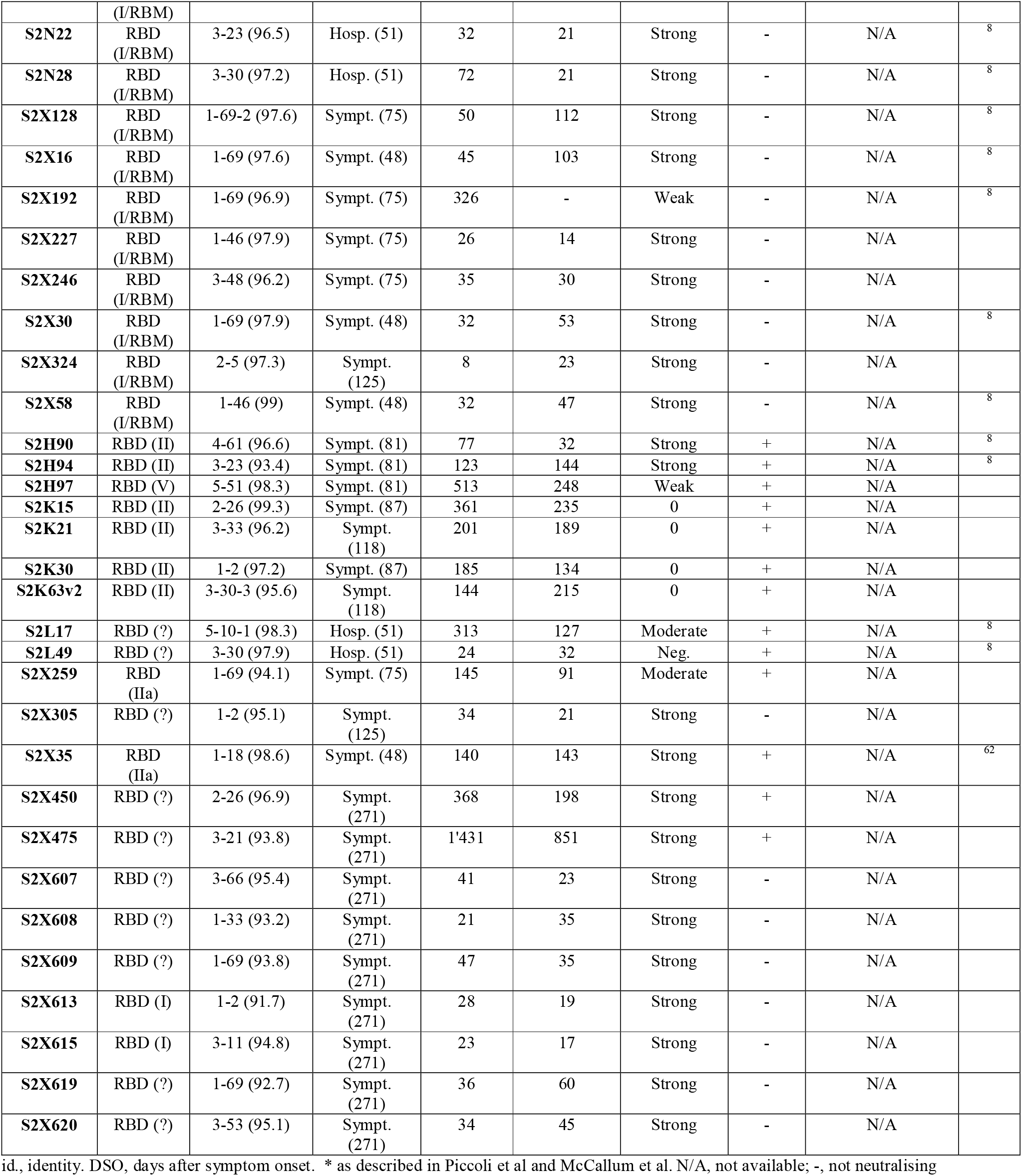
Neutralization, V gene usage and other properties of tested mAbs.

**The COVID-19 Genomics UK (COG-UK) Consortium**

**Funding acquisition, Leadership and supervision, Metadata curation, Project administration, Samples and logistics, Sequencing and analysis, Software and analysis tools, and Visualisation:**

Samuel C Robson ^13^.

**Funding acquisition, Leadership and supervision, Metadata curation, Project administration, Samples and logistics, Sequencing and analysis, and Software and analysis tools**:

Nicholas J Loman ^41^ and Thomas R Connor ^10,69^.

**Leadership and supervision, Metadata curation, Project administration, Samples and logistics, Sequencing and analysis, Software and analysis tools, and Visualisation:**

Tanya Golubchik ^5^.

**Funding acquisition, Metadata curation, Samples and logistics, Sequencing and analysis, Software and analysis tools, and Visualisation**:

Rocio T Martinez Nunez ^42^.

**Funding acquisition, Leadership and supervision, Metadata curation, Project administration, and Samples and logistics**:

Catherine Ludden ^88^.

**Funding acquisition, Leadership and supervision, Metadata curation, Samples and logistics, and Sequencing and analysis**:

Sally Corden ^69^.

**Funding acquisition, Leadership and supervision, Project administration, Samples and logistics, and Sequencing and analysis**:

Ian Johnston ^99^ and David Bonsall ^5^.

**Funding acquisition, Leadership and supervision, Sequencing and analysis, Software and analysis tools, and Visualisation:**

Colin P Smith ^87^ and Ali R Awan ^28^.

**Funding acquisition, Samples and logistics, Sequencing and analysis, Software and analysis tools, and Visualisation:**

Giselda Bucca ^87^.

**Leadership and supervision, Metadata curation, Project administration, Samples and logistics, and Sequencing and analysis:**

M. Estee Torok ^22^, ^101^.

**Leadership and supervision, Metadata curation, Project administration, Samples and logistics, and Visualisation:**

Kordo Saeed ^>81^, ^110^ and Jacqui A Prieto ^83^, ^109^.

**Leadership and supervision, Metadata curation, Project administration, Sequencing and analysis, and Software and analysis tools**:

David K Jackson ^99^.

**Metadata curation, Project administration, Samples and logistics, Sequencing and analysis, and Software and analysis tools:**

William L Hamilton ^22^.

**Metadata curation, Project administration, Samples and logistics, Sequencing and analysis, and Visualisation:**

Luke B Snell ^11^.

**Funding acquisition, Leadership and supervision, Metadata curation, and Samples and logistics:**

Catherine Moore ^69^.

**Funding acquisition, Leadership and supervision, Project administration, and Samples and logistics:**

Ewan M Harrison ^99^, ^88^.

**Leadership and supervision, Metadata curation, Project administration, and Samples and logistics:**

Sonia Goncalves ^99^.

**Leadership and supervision, Metadata curation, Samples and logistics, and Sequencing and analysis:**

Ian G Goodfellow ^24^, Derek J Fairley ^3^, ^72^, Matthew W Loose ^18^ and Joanne Watkins ^69^.

**Leadership and supervision, Metadata curation, Samples and logistics, and Software and analysis tools:**

Rich Livett ^99^.

**Leadership and supervision, Metadata curation, Samples and logistics, and Visualisation: Samuel Moses** ^**25, 106**^.

**Leadership and supervision, Metadata curation, Sequencing and analysis, and Software and analysis tools:**

Roberto Amato ^99^, Sam Nicholls ^41^ and Matthew Bull ^69^.

**Leadership and supervision, Project administration, Samples and logistics, and Sequencing and analysis:**

Darren L Smith ^37^, ^58^, ^105^.

**Leadership and supervision, Sequencing and analysis, Software and analysis tools, and Visualisation:**

Jeff Barrett ^99^, David M Aanensen ^14^, ^114^.

**Metadata curation, Project administration, Samples and logistics, and Sequencing and analysis:**

Martin D Curran ^65^, Surendra Parmar ^65^, Dinesh Aggarwal ^95^, ^99^, ^64^ and James G Shepherd ^48^.

**Metadata curation, Project administration, Sequencing and analysis, and Software and analysis tools:**

Matthew D Parker ^93^.

**Metadata curation, Samples and logistics, Sequencing and analysis, and Visualisation: Sharon Glaysher** ^**61**^.

**Metadata curation, Sequencing and analysis, Software and analysis tools, and Visualisation:**

Matthew Bashton ^37^, ^58^, Anthony P Underwood ^14^, ^114^, Nicole Pacchiarini ^69^ and Katie F

Loveson ^77^.

**Project administration, Sequencing and analysis, Software and analysis tools, and Visualisation:**

Alessandro M Carabelli ^88^.

**Funding acquisition, Leadership and supervision, and Metadata curation:**

Kate E Templeton ^53, 90^.

**Funding acquisition, Leadership and supervision, and Project administration:**

Cordelia F Langford ^99^, John Sillitoe ^99^, Thushan I de Silva ^93^ and Dennis Wang ^93^.

**Funding acquisition, Leadership and supervision, and Sequencing and analysis:**

Dominic Kwiatkowski ^99, 107^, Andrew Rambaut ^90^, Justin O’Grady ^70, 89^ and Simon Cottrell ^69^.

**Leadership and supervision, Metadata curation, and Sequencing and analysis:**

Matthew T.G. Holden ^68^ and Emma C Thomson ^48^.

**Leadership and supervision, Project administration, and Samples and logistics:**

Husam Osman ^64^, ^36^, Monique Andersson ^59^, Anoop J Chauhan ^61^ and Mohammed O Hassan-Ibrahim ^6^.

**Leadership and supervision, Project administration, and Sequencing and analysis:**

Mara Lawniczak ^99^.

**Leadership and supervision, Samples and logistics, and Sequencing and analysis:**

Ravi Kumar Gupta ^88, 113^, Alex Alderton ^99^, Meera Chand ^66^, Chrystala Constantinidou ^94^, Meera Unnikrishnan ^94^, Alistair C Darby ^92^, Julian A Hiscox ^92^ and Steve Paterson ^92^.

**Leadership and supervision, Sequencing and analysis, and Software and analysis tools:**

Inigo Martincorena ^99^, David L Robertson ^48^, Erik M Volz ^39^, Andrew J Page ^70^ and Oliver G Pybus ^23^.

**Leadership and supervision, Sequencing and analysis, and Visualisation:**

Andrew R Bassett ^99^.

**Metadata curation, Project administration, and Samples and logistics**:

Cristina V Ariani ^99^, Michael H Spencer Chapman ^99^, ^88^, Kathy K Li ^48^, Rajiv N Shah ^48^, Natasha G Jesudason ^48^ and Yusri Taha ^50^.

**Metadata curation, Project administration, and Sequencing and analysis:**

Martin P McHugh ^53^ and Rebecca Dewar ^53^.

**Metadata curation, Samples and logistics, and Sequencing and analysis:**

Aminu S Jahun ^24^, Claire McMurray ^41^, Sarojini Pandey ^84^, James P McKenna ^3^, Andrew Nelson ^58, 105^, Gregory R Young ^37, 58^, Clare M McCann ^58, 105^ and Scott Elliott ^61^.

Metadata curation, Samples and logistics, and Visualisation: Hannah Lowe ^25^.

**Metadata curation, Sequencing and analysis, and Software and analysis tools:**

Ben Temperton ^91^, Sunando Roy ^82^, Anna Price ^10^, Sara Rey ^69^ and Matthew Wyles ^93^.

Metadata curation, Sequencing and analysis, and Visualisation: Stefan Rooke ^90^ and Sharif Shaaban ^68^.

Project administration, Samples and logistics, Sequencing and analysis: Mariateresa de Cesare ^98^.

Project administration, Samples and logistics, and Software and analysis tools: Laura Letchford ^99^.

Project administration, Samples and logistics, and Visualisation: Siona Silveira ^81^, Emanuela Pelosi ^81^ and Eleri Wilson-Davies ^81^.

**Samples and logistics, Sequencing and analysis, and Software and analysis tools:**

Myra Hosmillo ^24^.

**Sequencing and analysis, Software and analysis tools, and Visualisation**:

Áine O’Toole ^90^, Andrew R Hesketh ^87^, Richard Stark ^94^, Louis du Plessis ^23^, Chris Ruis ^88^, Helen Adams ^4^ and Yann Bourgeois ^76^.

**Funding acquisition, and Leadership and supervision**:

Stephen L Michell ^91^, Dimitris Gramatopoulos ^84^, ^112^, Jonathan Edgeworth ^12^, Judith Breuer ^30,^

^82^, John A Todd ^98^ and Christophe Fraser ^5^.

**Funding acquisition, and Project administration:**

David Buck ^98^ and Michaela John ^9^.

Leadership and supervision, and Metadata curation: Gemma L Kay ^70^.

**Leadership and supervision, and Project administration:**

Steve Palmer ^99^, Sharon J Peacock ^88^, ^64^ and David Heyburn ^69^.

**Leadership and supervision, and Samples and logistics:**

Danni Weldon ^99^, Esther Robinson ^64^, ^36^, Alan McNally ^41, 86^, Peter Muir ^64^, Ian B Vipond ^64^, John BoYes ^29^, Venkat Sivaprakasam ^46^, Tranprit Salluja ^75^, Samir Dervisevic ^54^ and Emma J Meader ^54^.

**Leadership and supervision, and Sequencing and analysis:**

Naomi R Park ^99^, Karen Oliver ^99^, Aaron R Jeffries ^91^, Sascha Ott ^94^, Ana da Silva Filipe ^48^, David A Simpson ^72^ and Chris Williams ^69^.

**Leadership and supervision, and Visualisation:** Jane AH Masoli ^73, 91^.

**Metadata curation, and Samples and logistics:** Bridget A Knight ^73, 91^, Christopher R Jones ^73, 91^, Cherian Koshy ^1^, Amy Ash ^1^, Anna Casey ^71^, Andrew Bosworth ^64, 36^, Liz Ratcliffe ^71^, Li Xu-McCrae ^36^, Hannah M Pymont ^64^, Stephanie Hutchings ^64^, Lisa Berry ^84^, Katie Jones ^84^, Fenella Halstead ^46^, Thomas Davis ^21^, Christopher Holmes ^16^, Miren Iturriza-Gomara ^92^, Anita O Lucaci ^92^, Paul Anthony Randell ^38, 104^, Alison Cox ^38, 104^, Pinglawathee Madona ^38, 104^, Kathryn Ann Harris ^30^, Julianne Rose Brown ^30^, Tabitha W Mahungu ^74^, Dianne Irish-Tavares ^74^, Tanzina Haque ^74^, Jennifer Hart ^74^, Eric Witele ^74^, Melisa Louise Fenton ^75^, Steven Liggett ^79^, Clive Graham ^56^, Emma Swindells ^57^, Jennifer Collins ^50^, Gary Eltringham ^50^, Sharon Campbell ^17^, Patrick C McClure ^97^, Gemma Clark ^15^, Tim J Sloan ^60^, Carl Jones ^15^ and Jessica Lynch ^2, 111^.

**Metadata curation, and Sequencing and analysis:** Ben Warne ^8^, Steven Leonard ^99^, Jillian Durham ^99^, Thomas Williams ^90^, Sam T Haldenby ^92^, Nathaniel Storey ^30^, Nabil-Fareed Alikhan ^70^, Nadine Holmes ^18^, Christopher Moore ^18^, Matthew Carlile ^18^, Malorie Perry ^69^, Noel Craine ^69^, Ronan A Lyons ^80^, Angela H Beckett ^13^, Salman Goudarzi ^77^, Christopher Fearn ^77^, Kate Cook ^77^, Hannah Dent ^77^ and Hannah Paul ^77^.

**Metadata curation, and Software and analysis tools:** Robert Davies^99^.

**Project administration, and Samples and logistics:** Beth Blane ^88^, Sophia T Girgis ^88^, Mathew A Beale ^99^, Katherine L Bellis ^99, 88^, Matthew J Dorman ^99^, Eleanor Drury ^99^, Leanne Kane ^99^, Sally Kay ^99^, Samantha McGuigan ^99^, Rachel Nelson ^99^, Liam Prestwood ^99^, Shavanthi Rajatileka ^99^, Rahul Batra ^12^, Rachel J Williams ^82^, Mark Kristiansen ^82^, Angie Green ^98^, Anita Justice ^59^, Adhyana I.K Mahanama ^81, 102^ and Buddhini Samaraweera ^81, 102^.

**Project administration, and Sequencing and analysis:** Nazreen F Hadjirin ^88^ and Joshua Quick ^41^.

**Project administration, and Software and analysis tools:** Radoslaw Poplawski ^41^.

**Samples and logistics, and Sequencing and analysis:** Leanne M Kermack ^88^, Nicola Reynolds ^7^, Grant Hall ^24^, Yasmin Chaudhry ^24^, Malte L Pinckert ^24^, Iliana Georgana ^24^, Robin J Moll ^99^, Alicia Thornton ^66^, Richard Myers ^66^, Joanne Stockton ^41^, Charlotte A Williams ^82^, Wen C Yew ^58^, Alexander J Trotter ^70^, Amy Trebes ^98^, George MacIntyre-Cockett ^98^, Alec Birchley ^69^, Alexander Adams ^69^, Amy Plimmer ^69^, Bree Gatica-Wilcox ^69^, Caoimhe McKerr ^69^, Ember Hilvers ^69^, Hannah Jones ^69^, Hibo Asad ^69^, Jason Coombes ^69^, Johnathan M Evans ^69^, Laia Fina ^69^, Lauren Gilbert ^69^, Lee Graham ^69^, Michelle Cronin ^69^, Sara Kumziene-SummerhaYes ^69^, Sarah Taylor ^69^, Sophie Jones ^69^, Danielle C Groves ^93^, Peijun Zhang ^93^, Marta Gallis ^93^ and Stavroula F Louka ^93^.

Samples and logistics, and Software and analysis tools: Igor Starinskij ^48^.

**Sequencing and analysis, and Software and analysis tools:** Chris J Illingworth ^47^, Chris Jackson ^47^, Marina Gourtovaia ^99^, Gerry Tonkin-Hill ^99^, Kevin Lewis ^99^, Jaime M Tovar-Corona ^99^, Keith James ^99^, Laura Baxter ^94^, Mohammad T. Alam ^94^, Richard J Orton ^48^, Joseph Hughes ^48^, Sreenu Vattipally ^48^, Manon Ragonnet-Cronin ^39^, Fabricia F. Nascimento ^39^, David Jorgensen ^39^, Olivia Boyd ^39^, Lily Geidelberg ^39^, Alex E Zarebski ^23^, Jayna Raghwani ^23^, Moritz UG Kraemer ^23^, Joel Southgate ^10, 69^, Benjamin B Lindsey ^93^ and Timothy M Freeman ^93^.

**Software and analysis tools, and Visualisation:** Jon-Paul Keatley ^99^, Joshua B Singer ^48^, Leonardo de Oliveira Martins ^70^, Corin A Yeats ^14^, Khalil Abudahab ^14, 114^, Ben EW Taylor ^14, 114^ and Mirko Menegazzo ^14^.

**Leadership and supervision:** John Danesh ^99^, Wendy Hogsden ^46^, Sahar Eldirdiri ^21^, Anita Kenyon ^21^, Jenifer Mason ^43^, Trevor I Robinson ^43^, Alison Holmes ^38, 103^, James Price ^38, 103^, John A Hartley ^82^, Tanya Curran

^3^, Alison E Mather ^70^, Giri Shankar ^69^, Rachel Jones ^69^, Robin Howe ^69^ and Sian Morgan ^9^.

**Metadata curation:** Elizabeth Wastenge ^53^, Michael R Chapman ^34, 88, 99^, Siddharth Mookerjee ^38, 103^, Rachael Stanley ^54^, Wendy Smith ^15^, Timothy Peto ^59^, David Eyre ^59^, Derrick Crook ^59^, Gabrielle Vernet ^33^, Christine Kitchen ^10^, Huw Gulliver ^10^, Ian Merrick ^10^, Martyn Guest ^10^, Robert Munn ^10^, Declan T Bradley ^63, 72^ and Tim Wyatt ^63^.

**Project administration:** Charlotte Beaver ^99^, Luke Foulser ^99^, Sophie Palmer ^88^, Carol M Churcher ^88^, Ellena Brooks ^88^, Kim S Smith ^88^, Katerina Galai ^88^, Georgina M McManus ^88^, Frances Bolt ^38, 103^, Francesc Coll ^19^, Lizzie Meadows ^70^, Stephen W Attwood ^23^, Alisha Davies ^69^, Elen De Lacy ^69^, Fatima Downing ^69^, Sue Edwards ^69^, Garry P Scarlett ^76^, Sarah Jeremiah ^83^ and Nikki Smith ^93^.

**Samples and logistics:** Danielle Leek ^88^, Sushmita Sridhar ^88, 99^, Sally Forrest ^88^, Claire Cormie ^88^, Harmeet K Gill ^88^, Joana Dias ^88^, Ellen E Higginson ^88^, Mailis Maes ^88^, Jamie Young ^88^, Michelle Wantoch ^7^, Sanger Covid Team (www.sanger.ac.uk/covid-team) ^99^, Dorota Jamrozy ^99^, Stephanie Lo ^99^, Minal Patel ^99^, Verity Hill ^90^, Claire M Bewshea ^91^, Sian Ellard ^73, 91^, Cressida Auckland ^73^, Ian Harrison ^66^, Chloe Bishop ^66^, Vicki Chalker ^66^, Alex Richter ^85^, Andrew Beggs ^85^, Angus Best ^86^, Benita Percival ^86^, Jeremy Mirza ^86^, Oliver Megram ^86^, Megan Mayhew ^86^, Liam Crawford ^86^, Fiona Ashcroft ^86^, Emma Moles-Garcia ^86^, Nicola Cumley ^86^, Richard Hopes ^64^, Patawee Asamaphan ^48^, Marc O Niebel ^48^, Rory N Gunson ^100^, Amanda Bradley ^52^, Alasdair Maclean ^52^, Guy Mollett ^52^, Rachel Blacow ^52^, Paul Bird ^16^, Thomas Helmer ^16^, Karlie Fallon ^16^, Julian Tang ^16^, Antony D Hale ^49^, Louissa R Macfarlane-Smith ^49^, Katherine L Harper ^49^, Holli Carden ^49^, Nicholas W Machin ^45, 64^, Kathryn A Jackson ^92^, Shazaad S Y Ahmad ^45, 64^, Ryan P George ^45^, Lance Turtle ^92^, Elaine O’Toole ^43^, Joanne Watts ^43^, Cassie Breen ^43^, Angela Cowell ^43^, Adela Alcolea-Medina ^32, 96^, Themoula Charalampous ^12, 42^, Amita Patel ^11^, Lisa J Levett ^35^, Judith Heaney ^35^, Aileen Rowan ^39^, Graham P Taylor ^39^, Divya Shah ^30^, Laura Atkinson ^30^, Jack CD Lee

^30^, Adam P Westhorpe ^82^, Riaz Jannoo ^82^, Helen L Lowe ^82^, Angeliki Karamani ^82^, Leah Ensell ^82^, Wendy Chatterton ^35^, Monika Pusok ^35^, Ashok Dadrah ^75^, Amanda Symmonds ^75^, Graciela Sluga ^44^, Zoltan Molnar ^72^, Paul Baker ^79^, Stephen Bonner ^79^, Sarah Essex ^79^, Edward Barton ^56^, Debra Padgett ^56^, Garren Scott ^56^, Jane Greenaway ^57^, Brendan AI Payne ^50^, Shirelle Burton-Fanning ^50^, Sheila Waugh ^50^, Veena Raviprakash ^17^, Nicola Sheriff ^17^, Victoria Blakey^17^, Lesley-Anne Williams ^17^, Jonathan Moore ^27^, Susanne Stonehouse ^27^, Louise Smith ^55^, Rose

K Davidson ^89^, Luke Bedford ^26^, Lindsay Coupland ^54^, Victoria Wright ^18^, Joseph G Chappell ^97^, Theocharis Tsoleridis ^97^, Jonathan Ball ^97^, Manjinder Khakh ^15^, Vicki M Fleming ^15^, Michelle M Lister ^15^, Hannah C Howson-Wells ^15^, Louise Berry ^15^, Tim Boswell ^15^, Amelia Joseph ^15^, Iona Willingham ^15^, Nichola Duckworth ^60^, Sarah Walsh ^60^, Emma Wise ^2, 111^, Nathan Moore ^2, 111^, Matilde Mori ^2, 108, 111^, Nick Cortes ^2, 111^, Stephen Kidd ^2, 111^, Rebecca Williams ^33^, Laura Gifford ^69^, Kelly Bicknell ^61^, Sarah Wyllie ^61^, Allyson Lloyd ^61^, Robert Impey ^61^, Cassandra S Malone ^6^, Benjamin J Cogger ^6^, Nick Levene ^62^, Lynn Monaghan ^62^, Alexander J Keeley ^93^, David G Partridge ^78, 93^, Mohammad Raza ^78, 93^, Cariad Evans ^78, 93^ and Kate Johnson ^78, 93^.

**Sequencing and analysis:** Emma Betteridge ^99^, Ben W Farr ^99^, Scott Goodwin ^99^, Michael A Quail ^99^, Carol Scott ^99^, Lesley Shirley ^99^, Scott AJ Thurston ^99^, Diana Rajan ^99^, Iraad F Bronner ^99^, Louise Aigrain ^99^, Nicholas M Redshaw ^99^, Stefanie V Lensing ^99^, Shane McCarthy ^99^, Alex Makunin ^99^, Carlos E Balcazar ^90^, Michael D Gallagher ^90^, Kathleen A Williamson ^90^, Thomas D Stanton ^90^, Michelle L Michelsen ^91^, Joanna Warwick-Dugdale ^91^, Robin Manley ^91^, Audrey Farbos ^91^, James W Harrison ^91^, Christine M Sambles ^91^, David J Studholme ^91^, Angie Lackenby ^66^, Tamyo Mbisa ^66^, Steven Platt ^66^, Shahjahan Miah ^66^, David Bibby ^66^, Carmen Manso ^66^, Jonathan Hubb ^66^, Gavin Dabrera ^66^, Mary Ramsay ^66^, Daniel Bradshaw ^66^, Ulf Schaefer ^66^, Natalie Groves ^66^, Eileen Gallagher ^66^, David Lee ^66^, David Williams ^66^, Nicholas Ellaby ^66^, Hassan Hartman ^66^, Nikos Manesis ^66^, Vineet Patel ^66^, Juan Ledesma ^67^, Katherine A Twohig ^67^, Elias Allara ^64, 88^, Clare Pearson ^64, 88^, Jeffrey K. J. Cheng ^94^, Hannah E. Bridgewater ^94^, Lucy R. Frost ^94^, Grace Taylor-Joyce ^94^, Paul E Brown ^94^, Lily Tong ^48^, Alice Broos ^48^, Daniel Mair ^48^, Jenna Nichols ^48^, Stephen N Carmichael ^48^, Katherine L Smollett ^40^, Kyriaki Nomikou ^48^, Elihu Aranday-Cortes ^48^, Natasha Johnson ^48^, Seema Nickbakhsh ^48, 68^, Edith E Vamos ^92^, Margaret Hughes ^92^, Lucille Rainbow ^92^, Richard Eccles ^92^, Charlotte Nelson ^92^, Mark Whitehead ^92^, Richard Gregory ^92^, Matthew Gemmell ^92^, Claudia Wierzbicki ^92^, Hermione J Webster ^92^, Chloe L Fisher ^28^, Adrian W Signell ^20^, Gilberto Betancor ^20^, Harry D Wilson ^20^, Gaia Nebbia ^12^, Flavia Flaviani ^31^, Alberto C Cerda ^96^, Tammy V Merrill ^96^, Rebekah E Wilson ^96^, Marius Cotic ^82^, Nadua Bayzid ^82^, Thomas Thompson ^72^, Erwan Acheson ^72^, Steven Rushton ^51^, Sarah O’Brien ^51^, David J Baker ^70^, Steven Rudder ^70^, Alp Aydin ^70^, Fei Sang ^18^, Johnny Debebe ^18^, Sarah Francois ^23^, Tetyana I Vasylyeva ^23^, Marina Escalera Zamudio ^23^, Bernardo Gutierrez ^23^, Angela Marchbank ^10^, Joshua Maksimovic ^9^, Karla Spellman ^9^, Kathryn McCluggage ^9^, Mari Morgan ^69^, Robert Beer ^9^, Safiah Afifi ^9^, Trudy Workman ^10^, William Fuller ^10^, Catherine Bresner ^10^, Adrienn Angyal ^93^, Luke R Green ^93^, Paul J Parsons ^93^, Rachel M Tucker ^93^, Rebecca Brown ^93^ and Max Whiteley ^93^.

**Software and analysis tools:** James Bonfield ^99^, Christoph Puethe ^99^, Andrew Whitwham ^99^, Jennifier Liddle ^99^, Will Rowe ^41^, Igor Siveroni ^39^, Thanh Le-Viet ^70^ and Amy Gaskin ^69^.

**Visualisation:** Rob Johnson ^39^.

**1** Barking, Havering and Redbridge University Hospitals NHS Trust, **2** Basingstoke Hospital, **3** Belfast Health & Social Care Trust, **4** Betsi Cadwaladr University Health Board, **5** Big Data Institute, Nuffield Department of Medicine, University of Oxford, **6** Brighton and Sussex University Hospitals NHS Trust, **7** Cambridge Stem Cell Institute, University of Cambridge, **8** Cambridge University Hospitals NHS Foundation Trust, **9** Cardiff and Vale University Health Board, **10** Cardiff University, **11** Centre for Clinical Infection & Diagnostics Research, St. Thomas’ Hospital and Kings College London, **12** Centre for Clinical Infection and Diagnostics Research, Department of Infectious Diseases, Guy’s and St Thomas’ NHS Foundation Trust, **13** Centre for Enzyme Innovation, University of Portsmouth (PORT), **14** Centre for Genomic Pathogen Surveillance, University of Oxford, **15** Clinical Microbiology Department, Queens Medical Centre, **16** Clinical Microbiology, University Hospitals of Leicester NHS Trust, **17** County Durham and Darlington NHS Foundation Trust, **18** Deep Seq, School of Life Sciences, Queens Medical Centre, University of Nottingham, **19** Department of Infection Biology, Faculty of Infectious & Tropical Diseases, London School of Hygiene & Tropical Medicine, **20** Department of Infectious Diseases, King’s College London, **21** Department of Microbiology, Kettering General Hospital, **22** Departments of Infectious Diseases and Microbiology, Cambridge University Hospitals NHS Foundation Trust; Cambridge, UK, **23** Department of Zoology, University of Oxford, **24** Division of Virology, Department of Pathology, University of Cambridge, **25** East Kent Hospitals University NHS Foundation Trust, **26** East Suffolk and North Essex NHS Foundation Trust, **27** Gateshead Health NHS Foundation Trust, **28** Genomics Innovation Unit, Guy’s and St. Thomas’ NHS Foundation Trust, **29** Gloucestershire Hospitals NHS Foundation Trust, **30** Great Ormond Street Hospital for Children NHS Foundation Trust, **31** Guy’s and St. Thomas’ BRC, **32** Guy’s and St. Thomas’ Hospitals, **33** Hampshire Hospitals NHS Foundation Trust, **34** Health Data Research UK Cambridge, **35** Health Services Laboratories, **36** Heartlands Hospital, Birmingham, **37** Hub for Biotechnology in the Built Environment, Northumbria University, **38** Imperial College Hospitals NHS Trust, **39** Imperial College London, **40** Institute of Biodiversity, Animal Health & Comparative Medicine, **41** Institute of Microbiology and Infection, University of Birmingham, **42** King’s College London, **43** Liverpool Clinical Laboratories, **44** Maidstone and Tunbridge Wells NHS Trust, **45** Manchester University NHS Foundation Trust, **46** Microbiology Department, Wye Valley NHS Trust, Hereford, **47** MRC Biostatistics Unit, University of Cambridge, **48** MRC-University of Glasgow Centre for Virus Research, **49** National Infection Service, PHE and Leeds Teaching Hospitals Trust, **50** Newcastle Hospitals NHS Foundation Trust, **51** Newcastle University, **52** NHS Greater Glasgow and Clyde, **53** NHS Lothian, **54** Norfolk and Norwich University Hospital, **55** Norfolk County Council, **56** North Cumbria Integrated Care NHS Foundation Trust, **57** North Tees and Hartlepool NHS Foundation Trust, **58** Northumbria University, **59** Oxford University Hospitals NHS Foundation Trust, **60** PathLinks, Northern Lincolnshire & Goole NHS Foundation Trust, **61** Portsmouth Hospitals University NHS Trust, **62** Princess Alexandra Hospital Microbiology Dept., **63** Public Health Agency, **64** Public Health England, **65** Public Health England, Clinical Microbiology and Public Health Laboratory, Cambridge, UK, **66** Public Health England, Colindale, **67** Public Health England, Colindale, **68** Public Health Scotland, **69** Public Health Wales NHS Trust, **70** Quadram Institute Bioscience, **71** Queen Elizabeth Hospital, **72** Queen’s University Belfast, **73** Royal Devon and Exeter NHS Foundation Trust, **74** Royal Free NHS Trust, **75** Sandwell and West Birmingham NHS Trust, **76** School of Biological Sciences, University of Portsmouth (PORT), **77** School of Pharmacy and Biomedical Sciences, University of Portsmouth (PORT), **78** Sheffield Teaching Hospitals, **79** South Tees Hospitals NHS Foundation Trust, **80** Swansea University, **81** University Hospitals Southampton NHS Foundation Trust, **82** University College London, **83** University Hospital Southampton NHS Foundation Trust, **84** University Hospitals Coventry and Warwickshire, **85** University of Birmingham, **86** University of Birmingham Turnkey Laboratory, **87** University of Brighton, **88** University of Cambridge, **89** University of East Anglia, **90** University of Edinburgh, **91** University of Exeter, **92** University of Liverpool, **93** University of Sheffield, **94** University of Warwick, **95** University of Cambridge, **96** Viapath, Guy’s and St Thomas’ NHS Foundation Trust, and King’s College Hospital NHS Foundation Trust, **97** Virology, School of Life Sciences, Queens Medical Centre, University of Nottingham, **98** Wellcome Centre for Human Genetics, Nuffield Department of Medicine, University of Oxford, **99** Wellcome Sanger Institute, **100** West of Scotland Specialist Virology Centre, NHS Greater Glasgow and Clyde, **101** Department of Medicine, University of Cambridge, **102** Ministry of Health, Sri Lanka, **103** NIHR Health Protection Research Unit in HCAI and AMR, Imperial College London, **104** North West London Pathology, **105** NU-OMICS, Northumbria University, **106** University of Kent, **107** University of Oxford, **108** University of Southampton, **109** University of Southampton School of Health Sciences, **110** University of Southampton School of Medicine, **111** University of Surrey, **112** Warwick Medical School and Institute of Precision Diagnostics, Pathology, UHCW NHS Trust, **113** Wellcome Africa Health Research Institute Durban

---

**Extended Data Figure 1:**
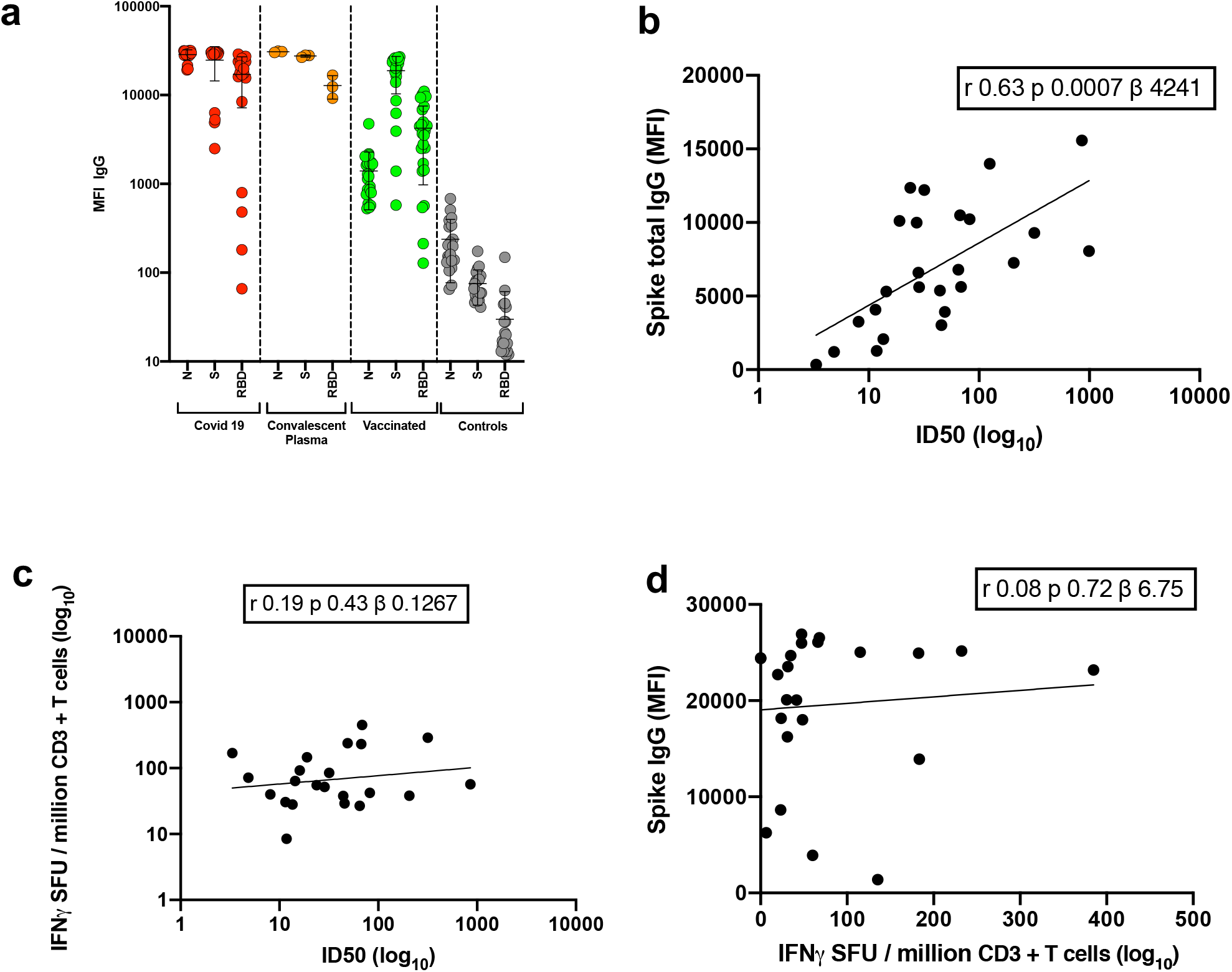
Immune responses three weeks after first dose of Pfizer SARS-CoV-2 vaccine BNT162b2. **a**, Serum IgG responses against N protein, Spike and the Spike Receptor Binding Domain (RBD) from first vaccine participants (green), recovered COVID-19 cases (red), 3 convalescent plasma units and healthy controls (grey) as measured by a flow cytometry based Luminex assay. MFI, mean fluorescence intensity. Geometric mean titre (GMT with standard deviation (s.d) of two technical repeats presented. **b**, Relationship between serum IgG responses as measured by flow cytometry and serum neutralisation ID50. **c**, Relationship between serum neutralisation ID50 and T cell responses against SARS-CoV-2 by IFN gamma ELISpot. SFU: spot forming units. **d**, Relationship between serum IgG responses and T cell responses. Simple linear regression is presented with Pearson correlation (r), P-value (p) and regression coefficient/slope (β).

**Extended Data Fig 2.**
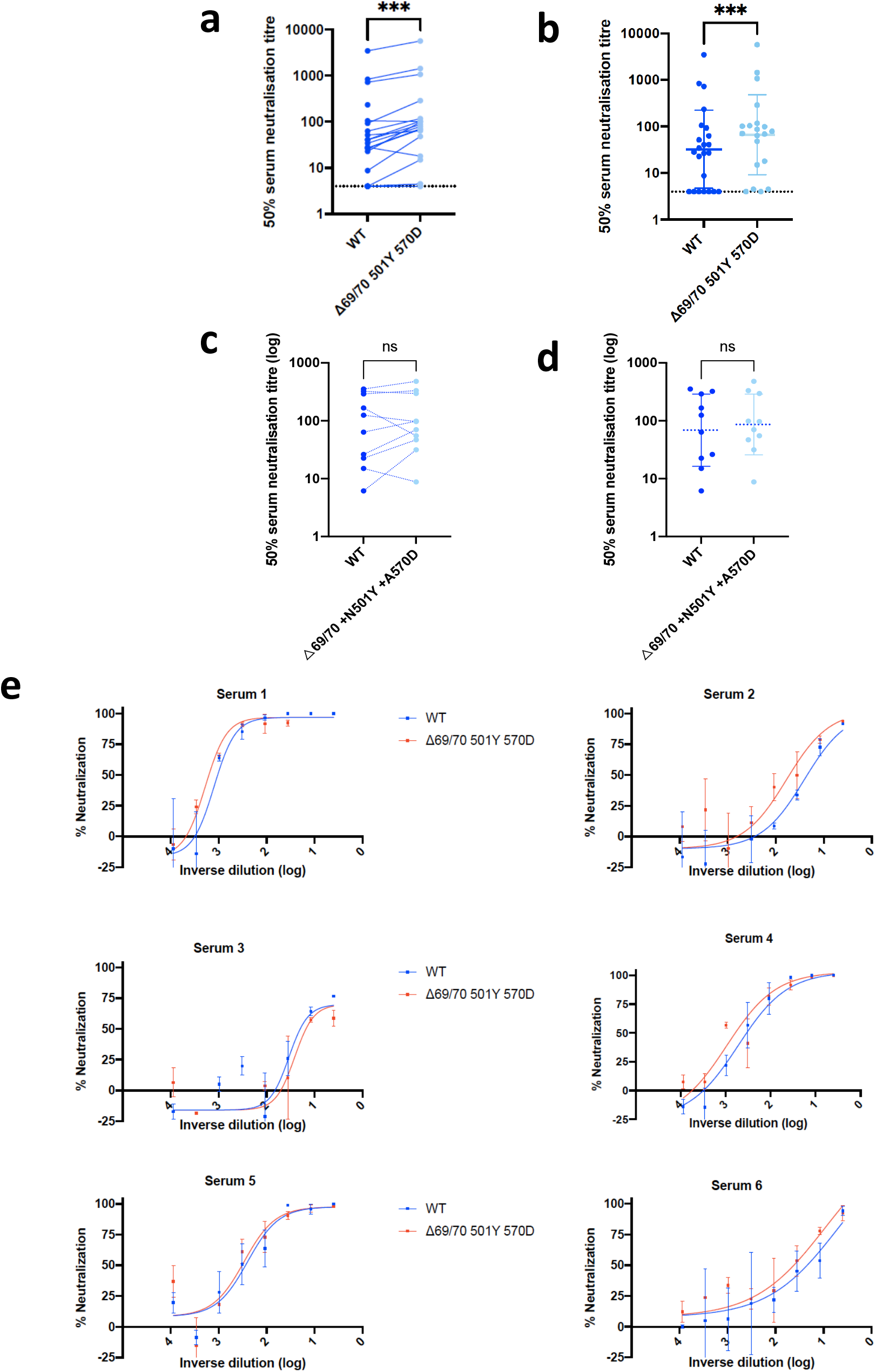
Neutralization by first dose BNT162b2 vaccine and convalescent sera against wild type and mutant (N501Y, A570D, ΔH69/V70) SARS-CoV-2 pseudotyped viruses: (**a-b**) Vaccine sera dilution for 50% neutralization against WT and Spike mutant with N501Y, A570D, ΔH69/V70. Geometric mean titre (GMT) + s.d of two independent experiments with two technical repeats presented. (**c-d**) Convalescent sera dilution for 50% neutralization against WT and Spike mutant with N501Y, A570D, ΔH69/V70. GMT + s.d of representative experiment with two technical repeats presented. **e**, Representative curves of convalescent serum log10 inverse dilution against % neutralization for WT v N501Y, A570D, ΔH69/V70. Where a curve is shifted to the right this indicates the virus is less sensitive to the neutralizing antibodies in the serum. Data are means of technical replicates and error bars represent standard error of the mean. Data are representative of 2 independent experiments. Limit of detection for 50% neutralization set at 10.

**Extended Data Fig 3.**
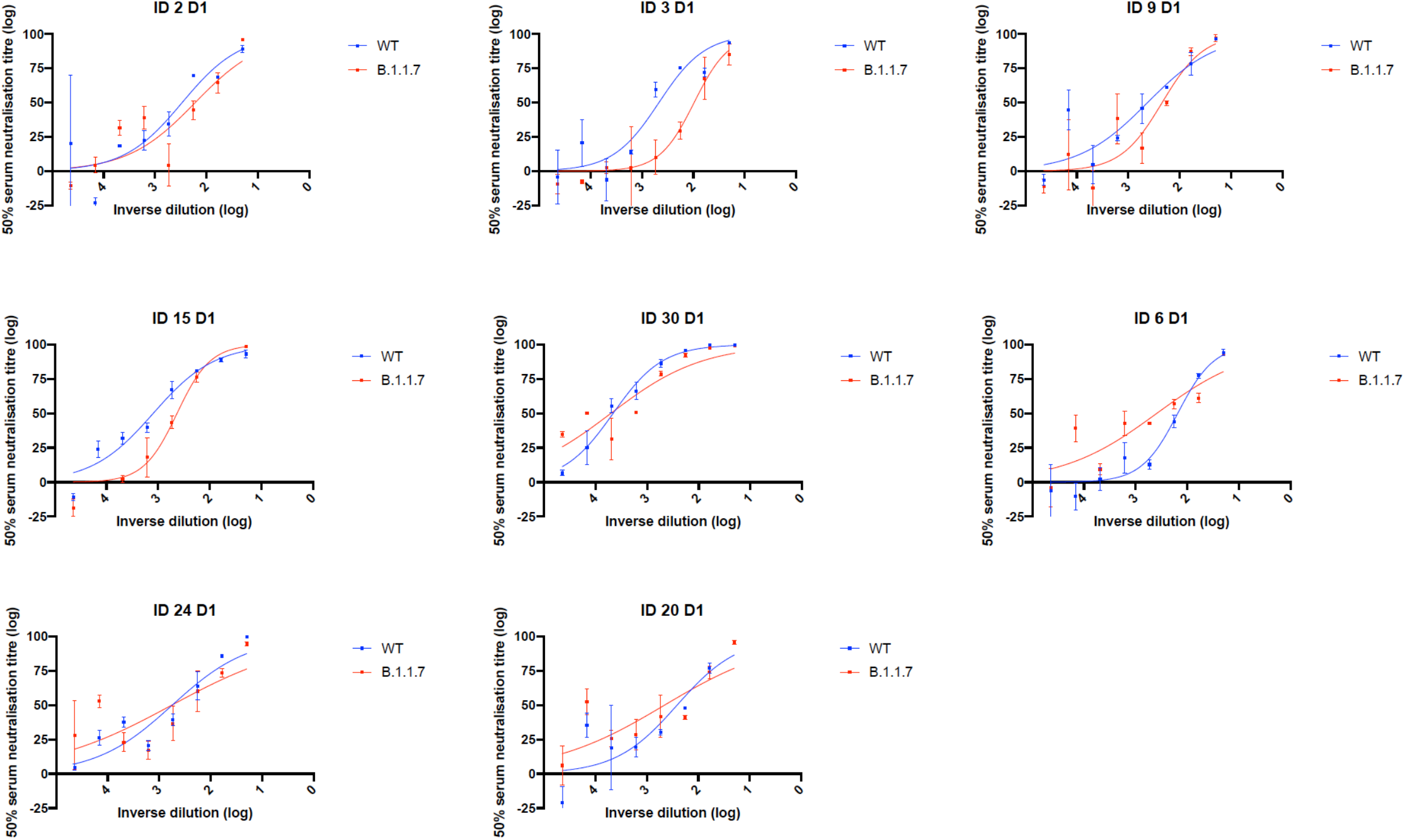
Representative neutralization curves of BNT162b2 vaccine sera against pseudovirus virus bearing eight Spike mutations present in B.1.1.7 versus wild type (all In Spike D614G background). Indicated is serum log_10_ inverse dilution against % neutralization. Where a curve is shifted to the right this indicates the virus is less sensitive to the neutralizing antibodies in the serum. Data are for first dose of vaccine (D1). Data points represent means of technical replicates and error bars represent standard error of the mean. Limit of detection for 50% neutralization set at 10.

**Extended Data Fig 4.**
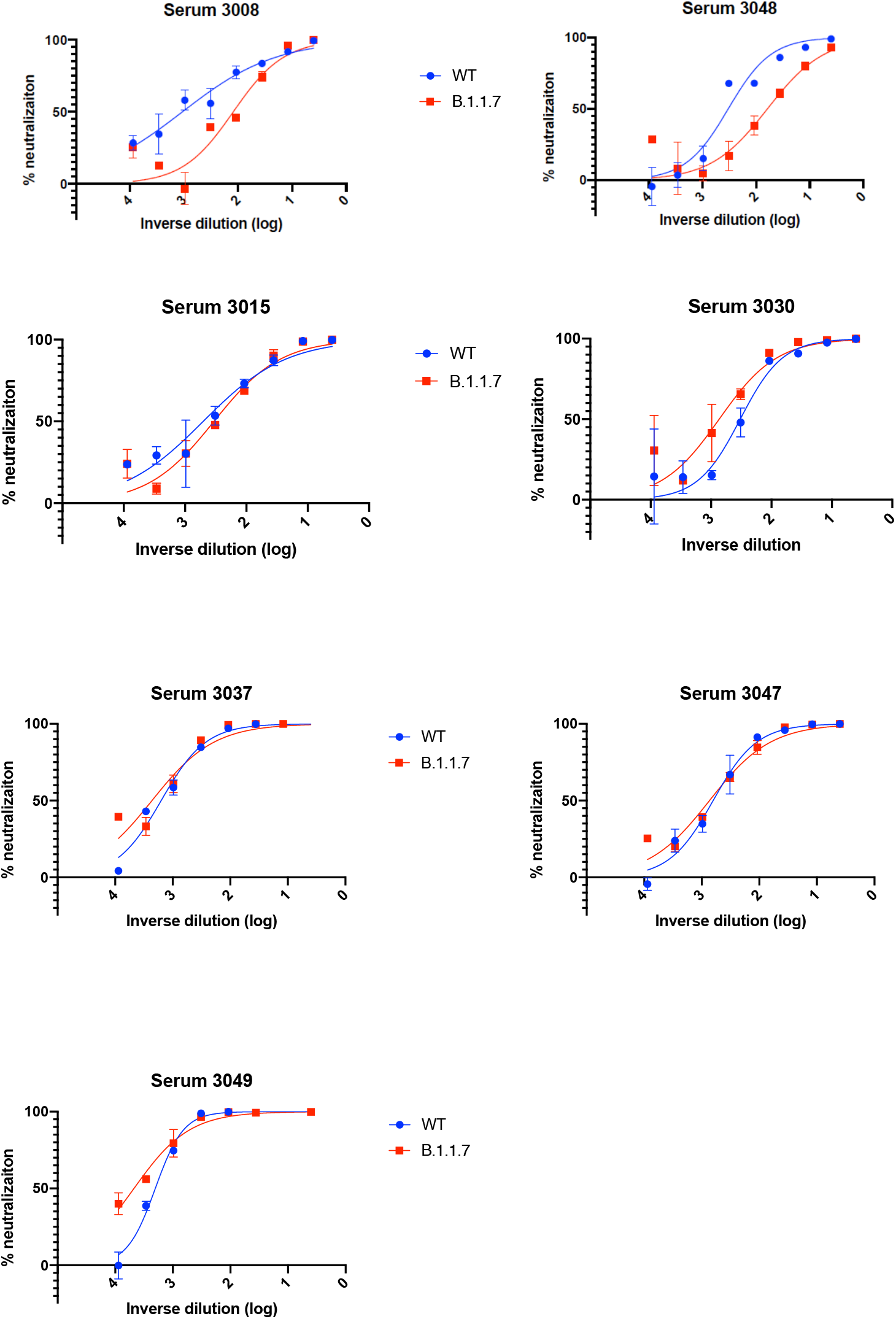
Representative neutralization curves of convalescent sera against wild type and B.1.1.7 Spike mutant SARS-CoV-2 pseudoviruses. Indicated is serum log_10_ inverse dilution against % neutralization. Where a curve is shifted to the right this indicates the virus is less sensitive to the neutralizing antibodies in the serum. Data points represent means of technical replicates and error bars represent standard error of the mean. Limit of detection for 50% neutralization set at 10.

**Extended Data Fig 5.**
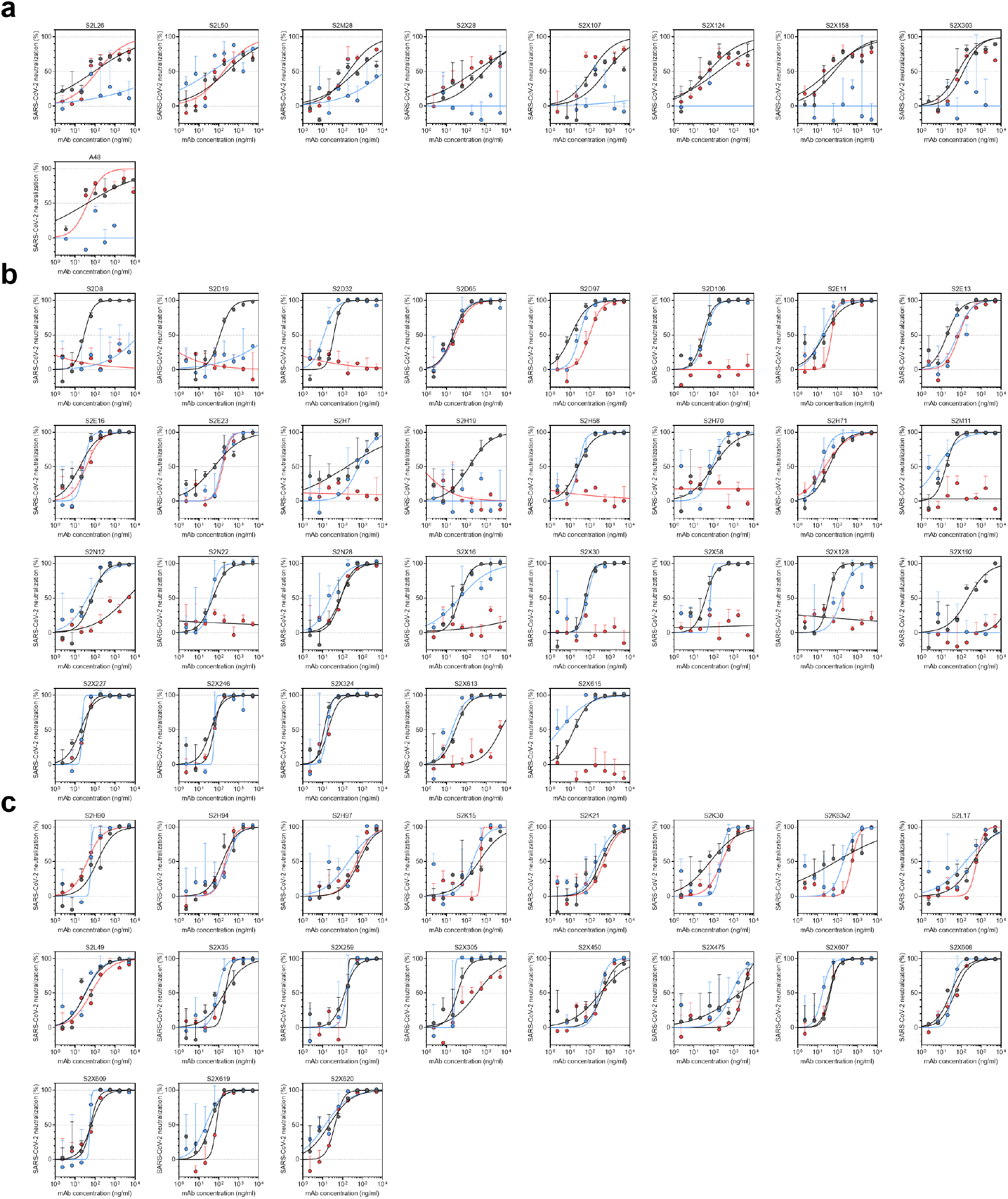
Neutralisation of WT (D614G), B.1.1.7 and TM (N501Y, E484K, K417N) SARS-CoV-2 Spike pseudotyped virus by a panel of 57 monoclonal antibodies (mAbs). **a-c**, Neutralisation of WT (black), B.1.1.7 (blue) and TM (red) SARS-CoV-2-MLV by 9 NTD-targeting (a), 29 RBM-targeting (b) and 19 non-RBM-targeting (c) mAbs.

**Extended Data Fig 6.**
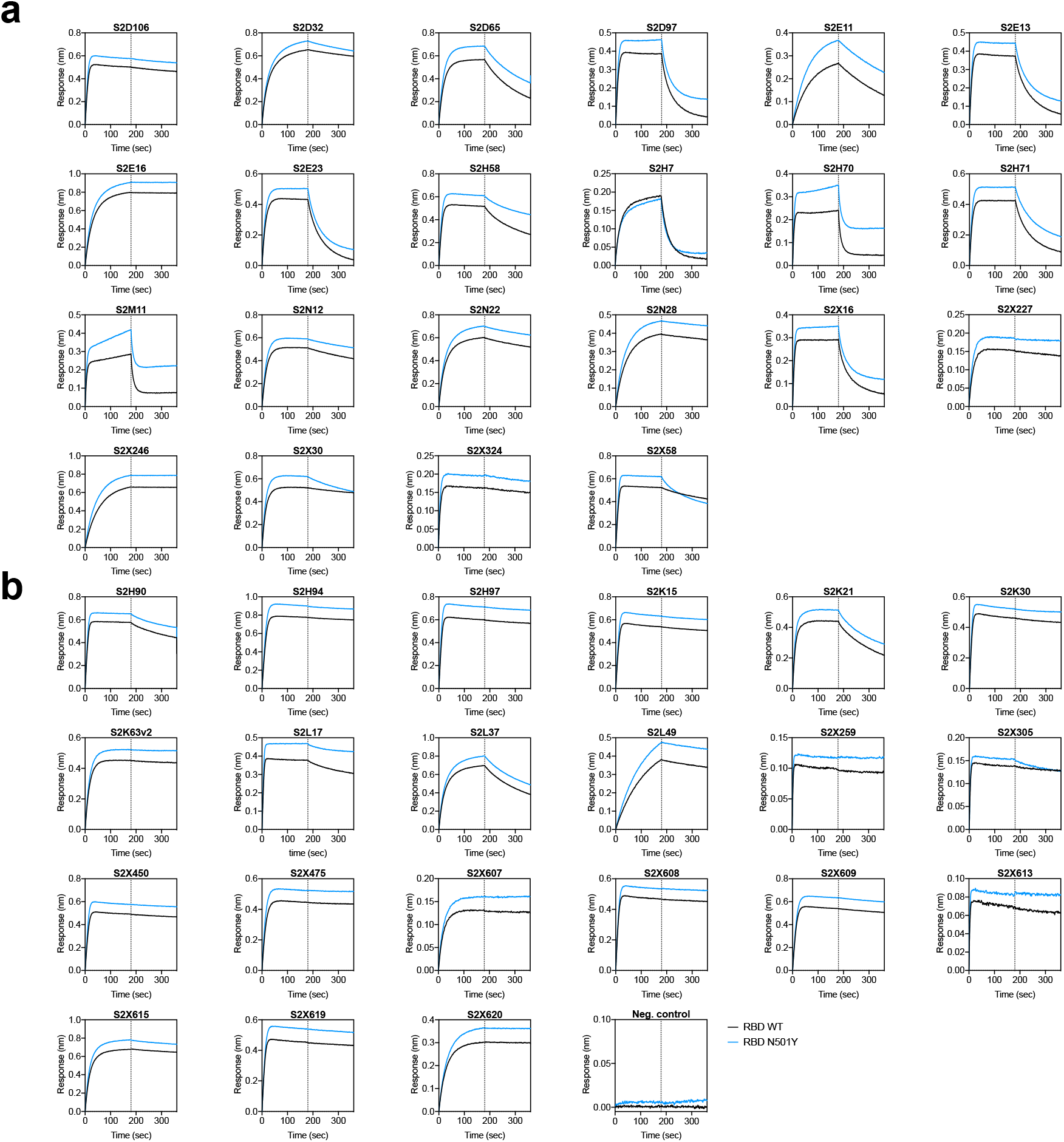
Kinetics of binding to WT and N501Y SARS-CoV-2 RBD of 43 RBD-specific mAbs. **a-b** Binding to WT (black) and N501Y (blue) RBD by 22 RBM-targeting (a) and 21 non-RBM-targeting (b) mAbs. An antibody of irrelevant specificity was included as negative control. mAbs: monoclonal antibodies

**Extended Data Fig 7.**
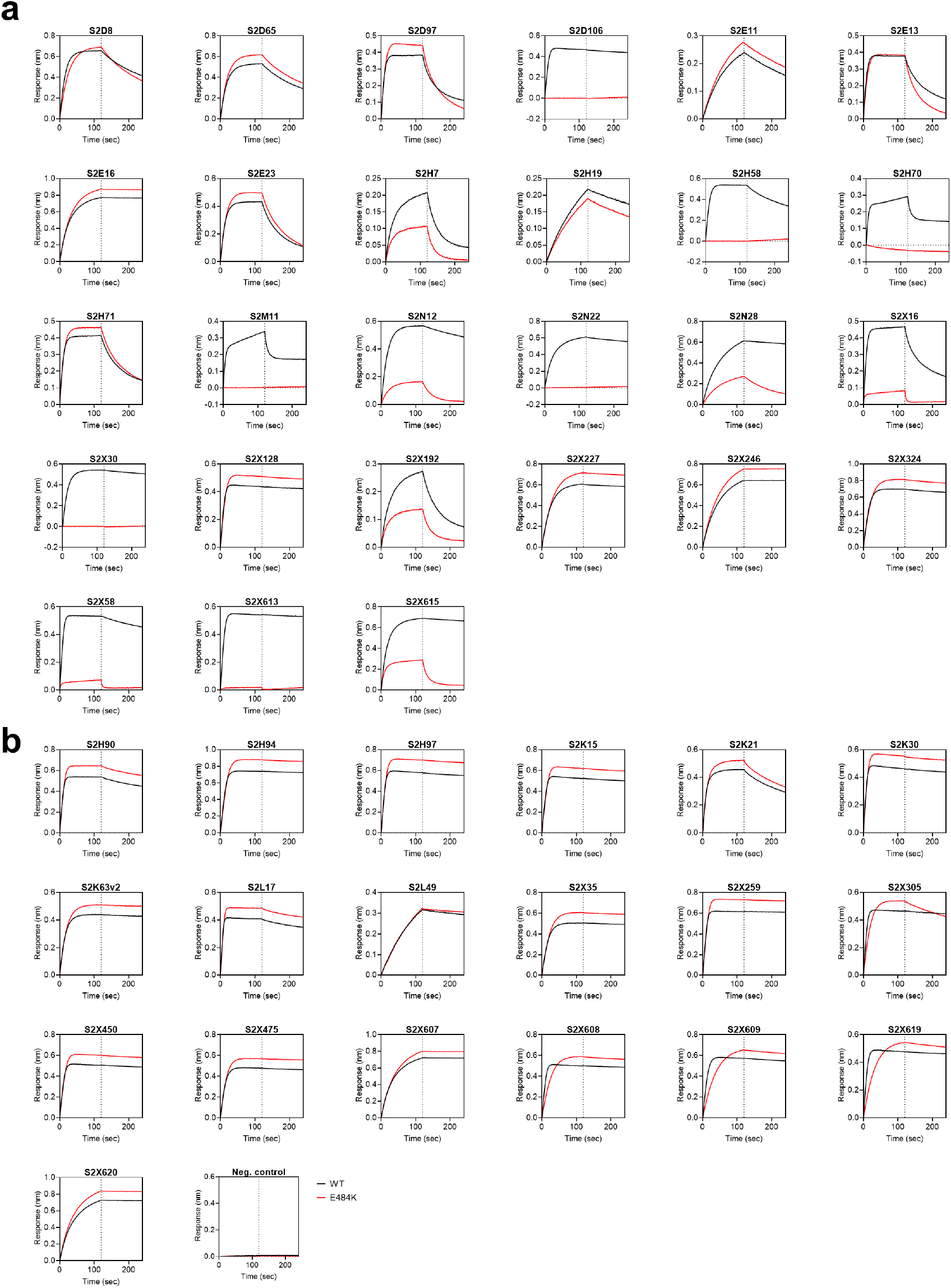
Kinetics of binding to WT and E484K SARS-CoV-2 RBD of 46 RBD-specific mAbs. **a-b**, Binding to WT (black) and E484K (red) RBD by 27 RBM-targeting (a) and 19 non-RBM-targeting (b) mAbs. An antibody of irrelevant specificity was included as negative control. mAbs: monoclonal antibodies

**Extended Data Fig 8.**
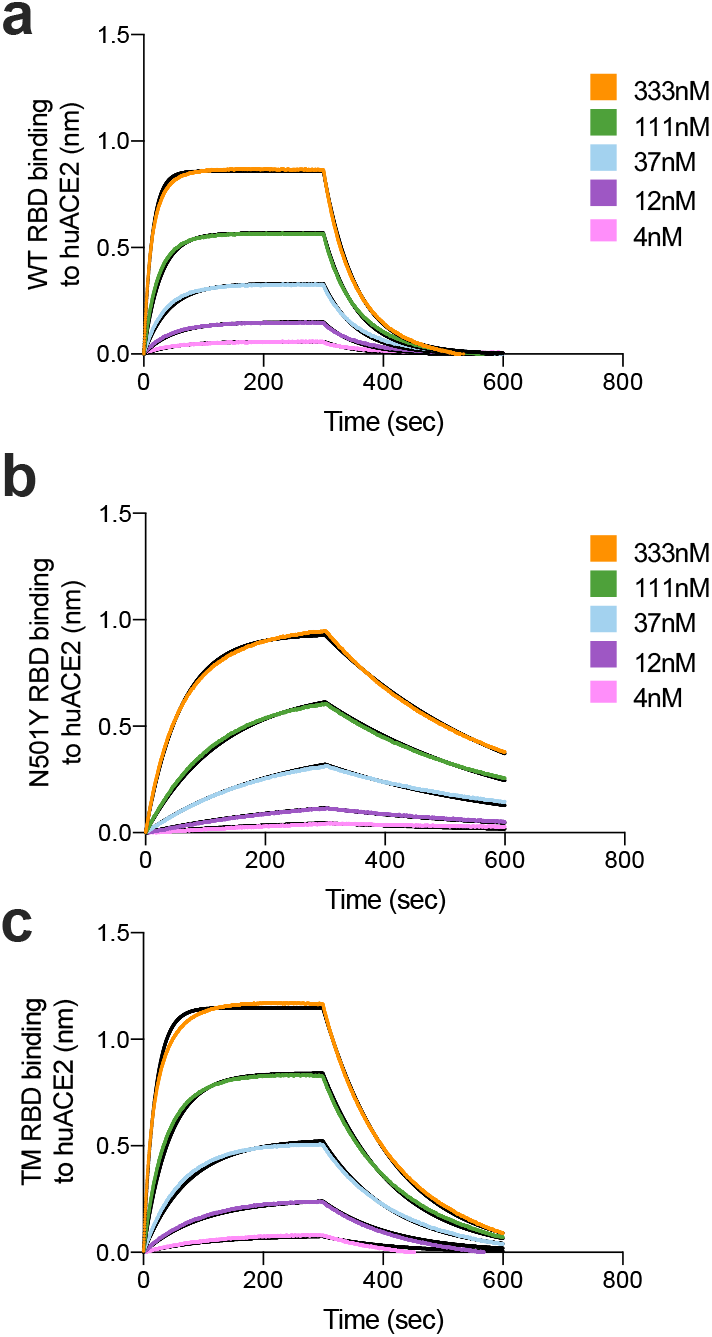
Binding of human ACE2 to SARS-CoV-2 WT, N501Y, TM (N501Y, E484K, K417N) RBDs. **a-b**. BLI binding analysis of the human ACE2 ectodomain (residues 1-615) to immobilized SARS-CoV-2 WT RBD (a) and B.1.1.7 RBD (c). Black lines correspond to a global fit of the data using a 1:1 binding model. RBD: receptor binding domain.

## References

1 Zhou, P. et al. A pneumonia outbreak associated with a new coronavirus of probable bat origin. Nature 579, 270–273, doi:10.1038/s41586-020-2012-7 (2020).

2 Davies, N. G. et al. Estimated transmissibility and severity of novel SARS-CoV-2 Variant of Concern 202012/01 in England. medRxiv, 2020.2012.2024.20248822, doi:10.1101/2020.12.24.20248822 (2020).

3 Volz, E. et al. Transmission of SARS-CoV-2 Lineage B.1.1.7 in England: Insights from linking epidemiological and genetic data. medRxiv, 2020.2012.2030.20249034, doi:10.1101/2020.12.30.20249034 (2021).

4 Korber, B. et al. Tracking Changes in SARS-CoV-2 Spike: Evidence that D614G Increases Infectivity of the COVID-19 Virus. Cell 182, 812–827 e819, doi:10.1016/j.cell.2020.06.043 (2020).

5 Yurkovetskiy, L. et al. Structural and Functional Analysis of the D614G SARS-CoV-2 Spike Protein Variant. Cell 183, 739–751 e738, doi:10.1016/j.cell.2020.09.032 (2020).

6 Martinot, M. et al. Remdesivir failure with SARS-CoV-2 RNA-dependent RNA-polymerase mutation in a B-cell immunodeficient patient with protracted Covid-19. Clin Infect Dis, doi:10.1093/cid/ciaa1474 (2020).

7 Kemp, S. et al. Neutralising antibodies in Spike mediated SARS-CoV-2 adaptation. medRxiv, 2020.2012.2005.20241927, doi:10.1101/2020.12.05.20241927 (2020).

8 Thomson, E. C. et al. The circulating SARS-CoV-2 spike variant N439K maintains fitness while evading antibody-mediated immunity. bioRxiv, 1–49, doi:papers3://publication/doi/10.1101/2020.11.04.355842 (2020).

9 Baden, L. R. et al. Efficacy and Safety of the mRNA-1273 SARS-CoV-2 Vaccine. N Engl J Med, doi:10.1056/NEJMoa2035389 (2020).

10 Polack, F. P. et al. Safety and Efficacy of the BNT162b2 mRNA Covid-19 Vaccine. N Engl J Med 383, 2603–2615, doi:10.1056/NEJMoa2034577 (2020).

11 Voysey, M. et al. Safety and efficacy of the ChAdOx1 nCoV-19 vaccine (AZD1222) against SARS-CoV-2: an interim analysis of four randomised controlled trials in Brazil, South Africa, and the UK. Lancet 397, 99–111, doi:10.1016/S0140-6736(20)32661-1 (2021).

12 Mulligan, M. J. et al. Phase I/II study of COVID-19 RNA vaccine BNT162b1 in adults. Nature 586, 589–593, doi:10.1038/s41586-020-2639-4 (2020).

13 Corbett, K. S. et al. SARS-CoV-2 mRNA Vaccine Development Enabled by Prototype Pathogen Preparedness. bioRxiv, 2020.2006.2011.145920, doi:10.1101/2020.06.11.145920 (2020).

14 Folegatti, P. M. et al. Safety and immunogenicity of the ChAdOx1 nCoV-19 vaccine against SARS-CoV-2: a preliminary report of a phase 1/2, single-blind, randomised controlled trial. Lancet 396, 467–478, doi:10.1016/S0140-6736(20)31604-4 (2020).

15 Kemp, S. A. et al. Recurrent emergence and transmission of a SARS-CoV-2 Spike deletion ΔH69/V70. bioRxiv, 2020.2012.2014.422555, doi:10.1101/2020.12.14.422555 (2020).

16 Tegally, H. et al. Emergence and rapid spread of a new severe acute respiratory syndrome-related coronavirus 2 (SARS-CoV-2) lineage with multiple spike mutations in South Africa. medRxiv, 2020.2012.2021.20248640, doi:10.1101/2020.12.21.20248640 (2020).

17 Faria, N. R. et al. Genomic characterisation of an emergent SARS-CoV-2 lineage in Manaus: preliminary findings, https://virological.org/t/genomic-characterisation-of-an-emergent-sars-cov-2-lineage-in-manaus-preliminary-findings/586 (2021).

18 Jackson, L. A. et al. An mRNA Vaccine against SARS-CoV-2 - Preliminary Report. N Engl J Med 383, 1920–1931, doi:10.1056/NEJMoa2022483 (2020).

19 Walsh, E. E. et al. Safety and Immunogenicity of Two RNA-Based Covid-19 Vaccine Candidates. New England Journal of Medicine 383, 2439–2450, doi:10.1056/NEJMoa2027906 (2020).

20 Schmidt, F. et al. Measuring SARS-CoV-2 neutralizing antibody activity using pseudotyped and chimeric viruses. 2020.2006.2008.140871, doi:10.1101/2020.06.08.140871 %J bioRxiv (2020).

21 Brouwer, P. J. M. et al. Potent neutralizing antibodies from COVID-19 patients define multiple targets of vulnerability. Science 369, 643–650, doi:10.1126/science.abc5902 (2020).

22 Wang, P. L. L; Iketani, S, Luo, Y; Guo, Y; Ho, D. Increased Resistance of SARS-CoV-2 Variants B.1.351 and B.1.1.7 to Antibody Neutralization. bioXriv (2021).

23 PHE. Public Health England statement on Variant of Concern and new Variant Under Investigation, https://www.gov.uk/government/news/phe-statement-on-variant-of-concern-and-new-variant-under-investigation (2021).

24 McCallum, M. et al. N-terminal domain antigenic mapping reveals a site of vulnerability for SARS-CoV-2. bioRxiv, doi:10.1101/2021.01.14.426475 (2021).

25 Thomson, E. C. et al. Circulating SARS-CoV-2 spike N439K variants maintain fitness while evading antibody-mediated immunity. Cell, doi:10.1016/j.cell.2021.01.037 (2021).

26 Greaney, A. J. et al. Comprehensive mapping of mutations to the SARS-CoV-2 receptor-binding domain that affect recognition by polyclonal human serum antibodies. Cell host & microbe, doi:https://doi.org/10.1016/j.chom.2021.02.003 (2021).

27 Andreano, E. et al. SARS-CoV-2 escape <em>in vitro</em> from a highly neutralizing COVID-19 convalescent plasma. bioRxiv, 2020.2012.2028.424451, doi:10.1101/2020.12.28.424451 (2020).

28 Walls, A. C. et al. Structure, Function, and Antigenicity of the SARS-CoV-2 Spike Glycoprotein. Cell 181, 281-292.e286, doi:papers3://publication/doi/10.1016/j.cell.2020.02.058 (2020).

29 Guan, Y. et al. Isolation and characterization of viruses related to the SARS coronavirus from animals in southern China. Science 302, 276–278, doi:10.1126/science.1087139 (2003).

30 Li, W. et al. Efficient replication of severe acute respiratory syndrome coronavirus in mouse cells is limited by murine angiotensin-converting enzyme 2. J Virol 78, 11429–11433, doi:10.1128/JVI.78.20.11429-11433.2004 (2004).

31 Li, W. et al. Receptor and viral determinants of SARS-coronavirus adaptation to human ACE2. The EMBO journal 24, 1634–1643 (2005).

32 Starr, T. N. et al. Deep Mutational Scanning of SARS-CoV-2 Receptor Binding Domain Reveals Constraints on Folding and ACE2 Binding. Cell 182, 1295–1310 e1220, doi:10.1016/j.cell.2020.08.012 (2020).

33 Wang, Z. et al. mRNA vaccine-elicited antibodies to SARS-CoV-2 and circulating variants. Nature, doi:10.1038/s41586-021-03324-6 (2021).

34 Wibmer, C. K. et al. SARS-CoV-2 501Y.V2 escapes neutralization by South African COVID-19 donor plasma. bioRxiv, doi:10.1101/2021.01.18.427166 (2021).

35 Verschoor, C. P. et al. Microneutralization assay titres correlate with protection against seasonal influenza H1N1 and H3N2 in children. PloS one 10, e0131531, doi:10.1371/journal.pone.0131531 (2015).

36 Kulkarni, P. S., Hurwitz, J. L., Simoes, E. A. F. & Piedra, P. A. Establishing Correlates of Protection for Vaccine Development: Considerations for the Respiratory Syncytial Virus Vaccine Field. Viral Immunol 31, 195–203, doi:10.1089/vim.2017.0147 (2018).

37 Goddard, N. L., Cooke, M. C., Gupta, R. K. & Nguyen-Van-Tam, J. S. Timing of monoclonal antibody for seasonal RSV prophylaxis in the United Kingdom. Epidemiol Infect 135, 159–162, doi:10.1017/S0950268806006601 (2007).

38 Mercado, N. B. et al. Single-shot Ad26 vaccine protects against SARS-CoV-2 in rhesus macaques. Nature 586, 583–588, doi:10.1038/s41586-020-2607-z (2020).

39 McMahan, K. et al. Correlates of protection against SARS-CoV-2 in rhesus macaques. Nature, doi:10.1038/s41586-020-03041-6 (2020).

40 Rathnasinghe, R. et al. The N501Y mutation in SARS-CoV-2 spike leads to morbidity in obese and aged mice and is neutralized by convalescent and post-vaccination human sera. medRxiv, 2021.2001.2019.21249592, doi:10.1101/2021.01.19.21249592 (2021).

41 Wang, Z. et al. mRNA vaccine-elicited antibodies to SARS-CoV-2 and circulating variants. bioRxiv, doi:10.1101/2021.01.15.426911 (2021).

42 Muik, A. et al. Neutralization of SARS-CoV-2 lineage B.1.1.7 pseudovirus by BNT162b2 vaccine-elicited human sera. Science, doi:10.1126/science.abg6105 (2021).

43 Wu, K. et al. mRNA-1273 vaccine induces neutralizing antibodies against spike mutants from global SARS-CoV-2 variants. bioXriv, doi:10.1101/2021.01.25.427948 (2021).

44 Xie, X. et al. Neutralization of N501Y mutant SARS-CoV-2 by BNT162b2 vaccine-elicited sera. bioRxiv, 2021.2001.2007.425740, doi:10.1101/2021.01.07.425740 (2021).

45 Suryadevara, N. et al. Neutralizing and protective human monoclonal antibodies recognizing the N-terminal domain of the SARS-CoV-2 spike protein. bioRxiv, 2021.2001.2019.427324, doi:10.1101/2021.01.19.427324 (2021).

46 Soh, W. T. et al. The N-terminal domain of spike glycoprotein mediates SARS-CoV-2 infection by associating with L-SIGN and DC-SIGN. bioRxiv, 1–30, doi:papers3://publication/doi/10.1101/2020.11.05.369264 (2020).

47 Greaney, A. J. et al. Comprehensive mapping of mutations to the SARS-CoV-2 receptor-binding domain that affect recognition by polyclonal human serum antibodies. bioRxiv, 2020.2012.2031.425021, doi:10.1101/2020.12.31.425021 (2021).

48 Greaney, A. J. et al. Complete mapping of mutations to the SARS-CoV-2 spike receptor-binding domain that escape antibody recognition. Cell Host & Microbe (2020).

49 Weisblum, Y. et al. Escape from neutralizing antibodies by SARS-CoV-2 spike protein variants. Elife 9, e61312, doi:10.7554/eLife.61312 (2020).

50 Corti, D. et al. A neutralizing antibody selected from plasma cells that binds to group 1 and group 2 influenza A hemagglutinins. Science 333, 850–856, doi:10.1126/science.1205669 (2011).

51 Pinto, D. et al. Cross-neutralization of SARS-CoV-2 by a human monoclonal SARS-CoV antibody. Nature 583, 290–295, doi:10.1038/s41586-020-2349-y (2020).

52 Tortorici, M. A. et al. Ultrapotent human antibodies protect against SARS-CoV-2 challenge via multiple mechanisms. Science, doi:10.1126/science.abe3354 (2020).

53 Gregson, J. et al. HIV-1 viral load is elevated in individuals with reverse transcriptase mutation M184V/I during virological failure of first line antiretroviral therapy and is associated with compensatory mutation L74I. Journal of Infectious Diseases (2019).

54 Forloni, M., Liu, A. Y. & Wajapeyee, N. Creating Insertions or Deletions Using Overlap Extension Polymerase Chain Reaction (PCR) Mutagenesis. Cold Spring Harb Protoc 2018, doi:10.1101/pdb.prot097758 (2018).

55 Case, J. B. et al. Neutralizing Antibody and Soluble ACE2 Inhibition of a Replication-Competent VSV-SARS-CoV-2 and a Clinical Isolate of SARS-CoV-2. Cell Host Microbe 28, 475–485 e475, doi:10.1016/j.chom.2020.06.021 (2020).

56 Naldini, L., Blomer, U., Gage, F. H., Trono, D. & Verma, I. M. Efficient transfer, integration, and sustained long-term expression of the transgene in adult rat brains injected with a lentiviral vector. Proceedings of the National Academy of Sciences of the United States of America 93, 11382–11388 (1996).

57 Gupta, R. K. et al. Full length HIV-1 gag determines protease inhibitor susceptibility within in vitro assays. AIDS 24, 1651 (2010).

58 Mlcochova, P. et al. Combined point of care nucleic acid and antibody testing for SARS-CoV-2 following emergence of D614G Spike Variant. Cell Rep Med, 100099, doi:10.1016/j.xcrm.2020.100099 (2020).

59 Walls, A. C. et al. Elicitation of Potent Neutralizing Antibody Responses by Designed Protein Nanoparticle Vaccines for SARS-CoV-2. Cell 183, 1367–1382 e1317, doi:10.1016/j.cell.2020.10.043 (2020).

60 Chi, X. et al. A neutralizing human antibody binds to the N-terminal domain of the Spike protein of SARS-CoV-2. Science, eabc6952–6913, doi:papers3://publication/doi/10.1126/science.abc6952 (2020).

61 Tortorici, M. A. et al. Ultrapotent human antibodies protect against SARS-CoV-2 challenge via multiple mechanisms. Science 4, eabe3354–3316, doi:papers3://publication/doi/10.1126/science.abe3354 (2020).

62 Piccoli, L. et al. Mapping neutralizing and immunodominant sites on the SARS-CoV-2 spike receptor-binding domain by structure-guided high-resolution serology. Cell, 1–55, doi:papers3://publication/doi/10.1016/j.cell.2020.09.037 (2020).

